# Sleep, pain, and neurodegeneration: A Mendelian randomization study

**DOI:** 10.1101/2021.03.08.21253133

**Authors:** Sandeep Grover, International Age-related Macular Degeneration Consortium (IAMDGC), Manu Sharma

## Abstract

**Objective:** To examine whether sleep and pain-related traits have a causal effect on the risk of neurodegeneration.

**Design:** Two-sample Mendelian randomization using an inverse-variance weighted (IVW) estimate of the summary effect estimates.

**Setting:** Genetic data on sleep and pain-related traits and neurodegenerative disorders (NDD) from various cohorts comprising individuals predominantly of European ancestry.

**Participants:** Participants from the International Sleep Genetic Epidemiology Consortium (ISGEC), UK Biobank sleep and chronotype research group, International Genomics of Alzheimer’s patients (IGAP), project MinE, International Age-related Macular Degeneration Consortium (IAMDGC), International Multiple Sclerosis Genetics Consortium (IMSGC), International Parkinson’s Disease Genomics Consortium (IPDGC)

**Exposures:** Self-reported chronotype (CHR), morning preference (MP), insomnia symptoms (INS), sleep duration (SP), short sleep (SS), long sleep (LS), and multisite chronic pain (MCP)

**Main outcome measures:** Age-related macular degeneration (AMD), Alzheimer’s disease (AD), Amyotrophic lateral sclerosis (ALS), Multiple sclerosis (MS), and Parkinson’s disease (PD)

**Results:** We considered a threshold of P=0.00142 as significant accounting for multiple testing, and P<0.05 was considered to be suggestive evidence for a potential association. Using direct MR, MP was observed as the strongest risk factor for AMD (OR_IVW_ = 1.19, 95% CI 1.08, 1.32, P = 0.00073). We observed suggestive evidence of influence of different sleep traits on neurodegeneration: CHR on AMD (OR_IVW_ = 1.27, 95% CI 1.08, 1.49, P = 0.0034), SS on AD (OR_IVW_ 1.26, 95% CI 1.08, 1.46, P = 0.0044), and INS on ALS (OR_IVW_ 1.55, 95% CI 1.12, 2.14, P = 0.0123). The association of SS with AD was, however, lost after the exclusion of overlapping UKB samples. Using pain as exposure, our study failed to observe any role of pain in neurodegeneration. Results were largely robust to reverse causal analyses and sensitivity analyses accounting for horizontal pleiotropy.

**Conclusions:** Our study highlighted the role of morning preference as a risk factor for AMD and provided suggestive evidence of different sleep traits on a wide spectrum of neurodegenerative diseases.

## Introduction

Patients with neurodegenerative diseases (NDD) often experience changes in circadian rhythmic activities^1^ ^2^. It has been observed that such changes often appear at an early stage of neurodegeneration and might be even a risk factor in younger healthy adults^3–7^. Many of the NDD patients with circadian disruptions also complain of painful symptoms of variable origin and intensity^8^. While chronically painful conditions are often associated with sleep disturbances in healthy individuals, the relationship between sleep and pain in NDD is poorly understood ^9^ ^10^. Similar to sleep disturbances, it has also been suggested that pain could be present even in the early phase of the disease and could even worsen the other symptoms^11–14^. However, both sleep and pain could often be treated and thereby can help maintain a stable quality of life in the absence of any disease-modifying treatments for NDD^15^. A greater understanding of the etiological relationship between sleep, pain, and neurodegeneration could thereby enable better management of NDD.

It is well recognized that circadian dysfunction in old age is due to degeneration of the suprachiasmatic nucleus (SCN) in the anterior hypothalamus, directly connected to the light-sensing retina^16^. Several behavioral markers, including sleep timing, daytime sleepiness, and rest-activity rhythmicity, can robustly identify any such disruption in circadian rhythms. However, individuals with neurodegeneration exhibit severe circadian disruptions compared to healthy adults of the same age^1^ ^2^. Different NDDs further exhibit marked heterogeneity in the manifestation of such disruptions, which could be attributed to the loss of different neuronal subpopulations in the SCN.

Several studies have shown the adverse impact of sleep deprivation in NDDs. For example, sleep deprivation has been shown to increase Amyloid-β and tau pathology in the Alzheimers’ disease (AD) mouse model ^17^. Similarly, disruption of non-24-h light-dark cycles in mouse models of amyotrophic lateral sclerosis (ALS) and Parkinson’s disease (PD) is associated with increased glial activation and neuroinflammation ^18^ ^19^. Clinically, patients with AD often show sleep-wake rhythm disorder, and patients with PD show a reduction in the amplitude of the circadian rhythm^20^ ^21^. A limited number of longitudinal studies have demonstrated the potential influence of circadian disruptions on predisposition to AD, PD, and related markers of neurodegeneration^7^ ^22–24^. For example, a 17 years follow-up study in 11247 adults showed an increased risk of dementia in individuals rising late in the morning^24^. Similarly, an 11 years follow-up study in 2920 older men without PD demonstrated an association of long-day sleepiness with increased risk of PD^7^.

Similar to the involvement of specific brain regions directly influencing the circadian rhythms, several brain regions also referred to as pain matrix, have been shown to be activated during pain perception^25^ ^26^. The pain matrix comprises of primary (S1) and secondary (S2) somatosensory cortex, insula, anterior cingulate cortex (ACC), amygdala, prefrontal cortex (PFC), and thalamus further show differential activation during acute and chronic pain^27^. Aging is specifically known to increase the likelihood of chronic pain and may amplify the neurodegeneration process^28^ ^29^.

However, the sparse number of large longitudinal studies and clinical trials have limited our progress in understanding the relationship between sleep, pain, and onset or progression of neurodegeneration, necessitating the need for searching alternative approaches for judging the causality. Mendelian randomization (MR) is one such approach that relies on using instruments or proxy markers of risk factors of interest in one population to judge causality with outcome in an independent population^30^. Such an approach could thereby eliminate the need for the co- existence of markers of sleep and pain and NDD in the same population. MR relies on assumptions of a strong association of genetic instruments with the risk factor of interest and influence of genetic instrument on outcome only through risk factor^31^. The approach has been able to successfully replicate several previously known findings from observational studies, including the association of telomerase length with AD^32^, alcohol drinking with amyotrophic lateral sclerosis (ALS)^33^, high-density lipoprotein cholesterol (HDL-C) with age-related macular degeneration (AMD)^34^, vitamin D with multiple sclerosis (MS)^35^, smoking with PD^36^.

To date, sleep duration has been investigated for its unidirectional causal association with AD^37^. The study, however, showed a lack of any causal relationship. Furthermore, MR studies exploring the causal association of pain with neurodegeneration are lacking. Considering the highly varied role of various behavioral biomarkers of circadian rhythm on neurodegeneration and potential overlapping etiology of sleep and pain, we adopted a highly comprehensive approach by exploiting the availability of genetic instruments for various markers of circadian rhythm, mainly sleep duration (SD)^38^, short sleep (SS)^38^, long sleep (LS)^38^, chronotype (CHR)^39^, morning person (MP)^39^, insomnia (INS)^40^, and multisite chronic pain (MCP)^41^, and NDDs including AMD^42^, AD^43^ ^44^, ALS^45^, MS^46^, and PD^47^ ^48^ to dissect the bi-directional relationship between sleep, pain, and neurodegeneration using an MR approach.

## Methods

### Study design and identification of datasets

We employed a two-sample MR study design using summary estimates to examine the lifelong effect of sleep and pain-related traits on the risk of neurodegeneration in the European populations. We used the latest available discovery cohorts of meta-analyses of GWAS datasets in the literature. We identified single nucleotide polymorphisms (SNPs) that influence circadian rhythm-related traits, including CHR^39^, MP^39^, INS^40^, SD^38^, SS^38^, LS^38^, and MCP^41^. The details of GWAS datasets employed for the current study are further shown in **Table 1**. We adopted a cutoff of 5×10^−8^ to select the genetic instruments. Concerning the outcome datasets, we used the discovery cohort of a recent meta-analysis of GWAS on AMD^42^, AD^43^, ALS^45^, MS^46^, and PD^47^, as described in **Table 1**. We further used the same datasets to prioritize genetic instruments to conduct MR using different NDDs as exposures for checking reverse causality (as discussed later in the sensitivity analysis).

**Table 1.** Details of discovery GWAS datasets explored and prioritized instruments used for direct and reverse causal analysis in the present study. The direct analysis was done using PD as an outcome, and the reverse was done using sleep and pain-related traits as an outcome.

| S.No. | Phenotype | Source study | Maximum sample size | P | Number of analyzed SNPs | Number of significant SNPs | Number of significant SNPs (post-clumping) ($R^2 < 0.001$) | Average F-statistics (Median (Range)) | $R^2$ (%) |
| --- | --- | --- | --- | --- | --- | --- | --- | --- | --- |
| **Sleep related traits** |  |  |  |  |  |  |  |  |  |
| 1 | Sleep duration (SD) | Dashti et al. 2019 | 446118 | $5 \times 10^{-8}$ | 14661601 | 7926 | 74 | 34.7 (29.6-220.9) | 0.731% |
| 2 | Short sleep (SS) | Dashti et al. 2019 | 106192 cases/<br>305742 controls | $5 \times 10^{-8}$ | 14661601 | 859 | 26 | 34.1 (29.9-77.0) | 0.045% |
| 3 | Long sleep (LS) | Dashti et al. 2019 | 34184 cases/<br>305742 controls | $5 \times 10^{-8}$ | 14661601 | 3901 | 9 | 32.4 (29.9-53.0) | 0.006% |
| 4 | Chronotype (CHR) | Jones et al. 2019 | 449734 | $5 \times 10^{-8}$ | 11977111 | 15152 | 156 | 39.4 (28.2-209.4) | 2.683% |
| 5 | Morning person (MP) | Jones et al. 2019 | 252287 cases/<br>150908 controls | $5 \times 10^{-8}$ | 11977111 | 10949 | 127 | 37.9 (29.0-168.5) | 5.748% |
| 6 | Insomnia (INS) | Jansen et al. 2018 | 109389 cases/<br>277144 controls | $5 \times 10^{-8}$ | 10862567 | 463 | 13 | 34.4 (30.4-94.7) | 0.712% |
| **Pain related trait** |  |  |  |  |  |  |  |  |  |
| 1 | Multisite chronic pain (MCP) | Johnston et al. 2019 | 387649 | $5 \times 10^{-8}$ | 9926106 | 1746 | 41 | 34.1 (30.0-54.6) | 0.341% |
| **Disease trait** |  |  |  |  |  |  |  |  |  |
| 1 | Age-related macular degeneration (AMD) | Fritsche et al. 2016 | 16144 cases/<br>17832 controls | $5 \times 10^{-8}$ | 120,238,30 | 7218 | 42 | 47.5 (29.2-382.5) | NA |
| 2 | Alzheimer's disease (AD) | Jansen et al. 2018 | 71880 cases/<br>383,378 controls | $5 \times 10^{-8}$ | 33,672,99 | 2357 | 27 | 42.2 (30.2-422.5) | NA |
| 3 | Amyotrophic lateral sclerosis (ALS) | van Rheenen et al. 2016 | 12577 cases/<br>23475 controls | $5 \times 10^{-8}$ | 870, 945,2 | 125 | 4 | 37.2 (32.2-80.1) | NA |
| 4 | Multiple sclerosis (MS) | Patsopoulos et al. 2017 | 47351 cases/<br>68284 controls | $5 \times 10^{-8}$ | 859, 365,0 | 26403 | 74 | 41.9 (29.8-561.9) | NA |
| 5 | Parkinson's disease (PD) | Nalls et al. 2019 | 33674 cases,<br>449056 controls | $5 \times 10^{-8}$ | 175,137,73 | 3465 | 23 | 43.6 (30.0-181.5) | NA |
| 1 | Alzheimer's disease (AD) | Lambert et al. 2013 | 17008 cases/<br>37154 controls | $5 \times 10^{-8}$ | 7055881 | 1090 | 18 | 37.9 (29.7-82.4) | NA |
| 2 | Parkinson's disease (PD) | Nalls et al. 2014 | 9581 cases/<br>33245 controls | $5 \times 10^{-8}$ | 8543957 | 3209 | 9 | 49.8 (33.1-175.7) | NA |

### Prioritization of genetic variants

All the prioritized SNP IDs and positions were synchronized with the NCBI GRCh37 assembly. We further checked for the validity of MR assumptions by excluding SNPs with F-statistics >10 and loci known to be directly involved in neurodegeneration based on the existing evidence from previously published literature.

As the selected genetic instruments could be correlated, we performed clumping of the significantly associated SNPs in each GWAS dataset with the clump_data function of the TwoSampleMR Package (version 0.4.25) in R (version 3.6.1)^49^. We employed a clumping window of 10,000 kb and linkage disequilibrium (LD; i.e. r^2^) cutoff of 0.001 and used the European population in the 1000Genome Phase 3v5 dataset to identify the leading SNP^50^.

The leading SNPs were further checked for availability in the respective outcome datasets. If a specific SNP was not available, a proxy SNP (r^2^ > 0.8) was used, when possible. We further computed the pooled variance (R^2^) for the respective risk factor using effect estimates (β_x_) and effect allele frequencies (EAF) of individual genetic instruments, i.e. R^2^ = 2* ^2^*EAF*(1-EAF). The detectable risky and protective effect estimate at 80% power was computed for each neurodegenerative disorder as an outcome at various pooled variances explained by the genetic instruments (ranging from 0.25% to 7.5%) using the method described by Brion *et al.* (available at http://cnsgenomics.com/shiny/mRnd)51. To compute the effect estimates at specific variance for a given outcome, we employed a sample size of each outcome dataset, the proportion of patients in the same dataset, and a threshold p-value of 1.42 × 10^−3^ (see the section below).

### Causal effect estimation and test of pleiotropy

We used the inverse variance-weighted (IVW) effect method as the primary method to compute the causal effect estimates^52^. We computed the causal estimates as odds ratio (OR) per unit of standard deviation (SD) for continuous traits and ORs for the outcome per unit log-odds of categorical traits. We employed a conservative Bonferroni correction of the significance level to account for 35 independent tests, including forward and reverse MR (threshold p-value = 0.00142, i.e. 0.05/35). The heterogeneity was judged using the Cochrane Q-statistics and I^2^ for the IVW method along with Rucker’s Q-statistics and the Intercept deviation test for the MR-Egger’s method^31^ ^53–55^. We also used MR-PRESSO (Mendelian Randomization Pleiotropy RESidual Sum and Outlier) global test to evaluate horizontal pleiotropy^56^. Lastly, we performed an MR Steiger test of directionality to check the assumption whether an exposure causes outcome is valid using the TwoSampleMR Package (version 0.4.25) in R (version 3.6.1)^49^

### Sensitivity analysis

#### Multiple MR methods

Several approaches were employed to rule out the influence of potential pleiotropic variants on the overall results. We used multiple modern MR methods, including MR-Egger, weighted median (WME), weighted mode (MBE) methods to check the reliability of estimates^31^ ^54 57–59^.

#### Exclusion of overlapping samples

Since most of the recent meta-analyses of GWAS compute effect estimates by pooling UK Biobank (UKB) datasets with previously available datasets, and the existence of any overlapping samples in exposure and outcome datasets could bias the effect estimates towards the confounded observational estimates^60^, we also used the NDD datasets without UKB samples, when possible^44^ ^61^.

#### Reverse causality check

We conducted MR in the reverse direction to check to confirm the directionality of the observed associations.

#### Exclusion of potential pleiotropic variants

We further employed a leave-one-out and leave-one-group-out cross-validation approach to rule out the influence of outlier variants and variants known to be associated with confounders of the relationship between the respective exposure and outcome datasets. We specifically employed the Phenoscanner database (http://phenoscanner.medschl.cam.ac.uk) to identify such genetic variants known to be associated with potential confounders^62^. However, in the absence of knowledge of potential confounders, we adopted a more conservative approach, and all those genetic loci known to be associated with non-sleep-related traits were assumed to be pleiotropic loci. We identified such loci by searching for all the genetic variants in high LD with genetic instruments prioritized for the present study using r^2^ > 0.9 for previously reported associations in European populations. We also used visual approaches, including scatter plots and funnel plots, to identify the outlier variants.

#### Influence of regional brain expression

We further evaluated the potential biological influence of different brain regions in their respective contribution to the causal effect estimate by analyzing gene expression data for the available genetic variants from the Genotype-Tissue Expression Project (https://www.gtexportal.org)63. The identified genetic variants were then categorized as per their expression in specific brain regions and checked for their influence on the causal estimate using a leave-one-group-out cross-validation approach.

## Results

### Study design and identification of datasets

The genetic instruments were identified that influence sleep and pain-related traits through the latest publicly available meta-analysis of GWAS summary datasets (**Table 1**). Concerning sample size, GWAS on all sleep and pain-related traits employed approximately 0.4 million individuals except SS and LS with a low sample size of 106,192 and 34,184. On the other hand, among NDDs, only AD and PD datasets had a similar sample size with 0.45 million individuals, comprising 71,880 and 33,674 cases. Concerning depth of genomic coverage, GWAS for various sleep and pain-related traits showed fairly uniform coverage with 0.9-1.5 million SNPs covering the whole genome in different summary datasets. Concerning NDDs, genomes of PD patients were best covered with the availability of data on up to 1.75 million SNPs.

### Prioritization of genetic variants

Concerning sleep and pain-related traits, the number of independent genetic instruments identified varied from 9 for LS to 156 for MP. Among NDDs, only four genetic instruments were identified for ALS. On the other hand, a total of 74 SNPs were representative of MS. We further observed that none of the genetic instruments had an F-statistic <10, thereby validating MR Assumption I. Overall, we identified 771 genetic instruments to check the bidirectional causality check between sleep, pain, and neurodegeneration with F-statistic for individuals SNPs ranging from 28.2 to 422.5. The detectable effect estimates for different NDDs as outcomes at 80% power and a type 1 error rate of 1.42 × 10^−3^ are further shown in **Supplementary Table 1**.

### Causal effect estimation

Data used for computation of causal effect estimates have been further provided in **Supplementary Table 2**. The causal effect estimates using various MR approaches and heterogeneity analysis measures used to judge the robustness of the estimates have been further provided in **Table 2** for the direct causal estimates for NDDs as outcomes.

**Table 2.**
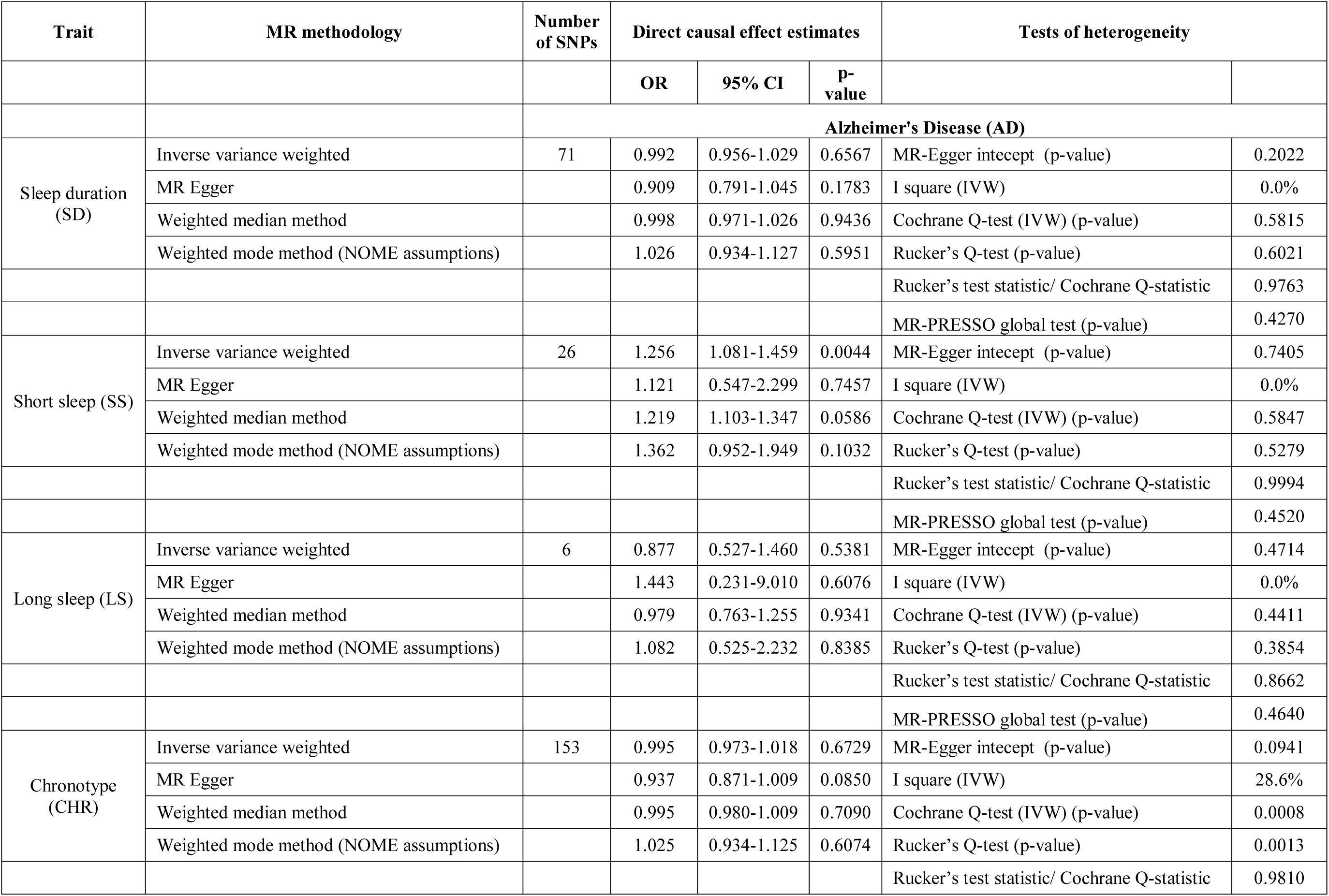
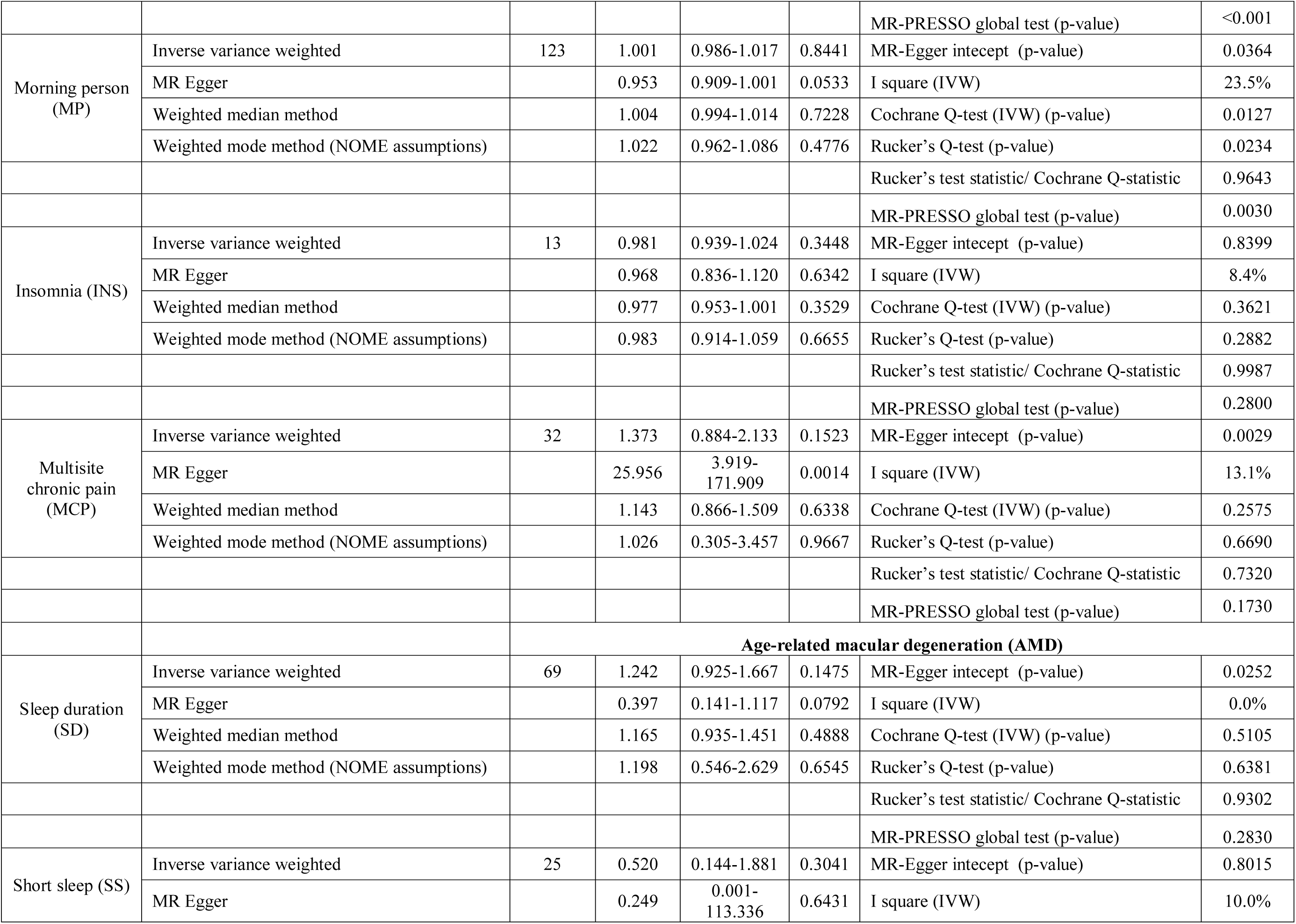
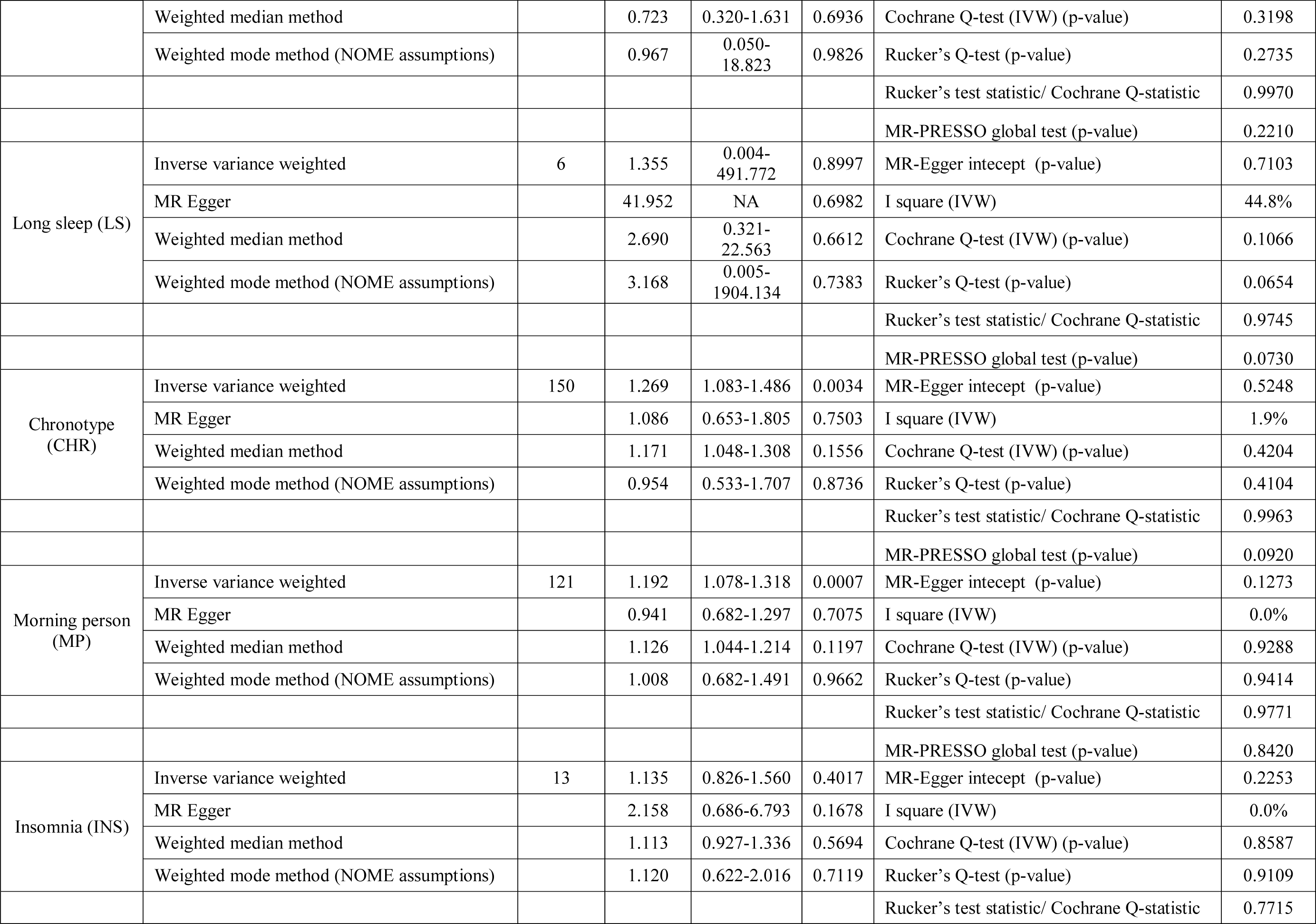
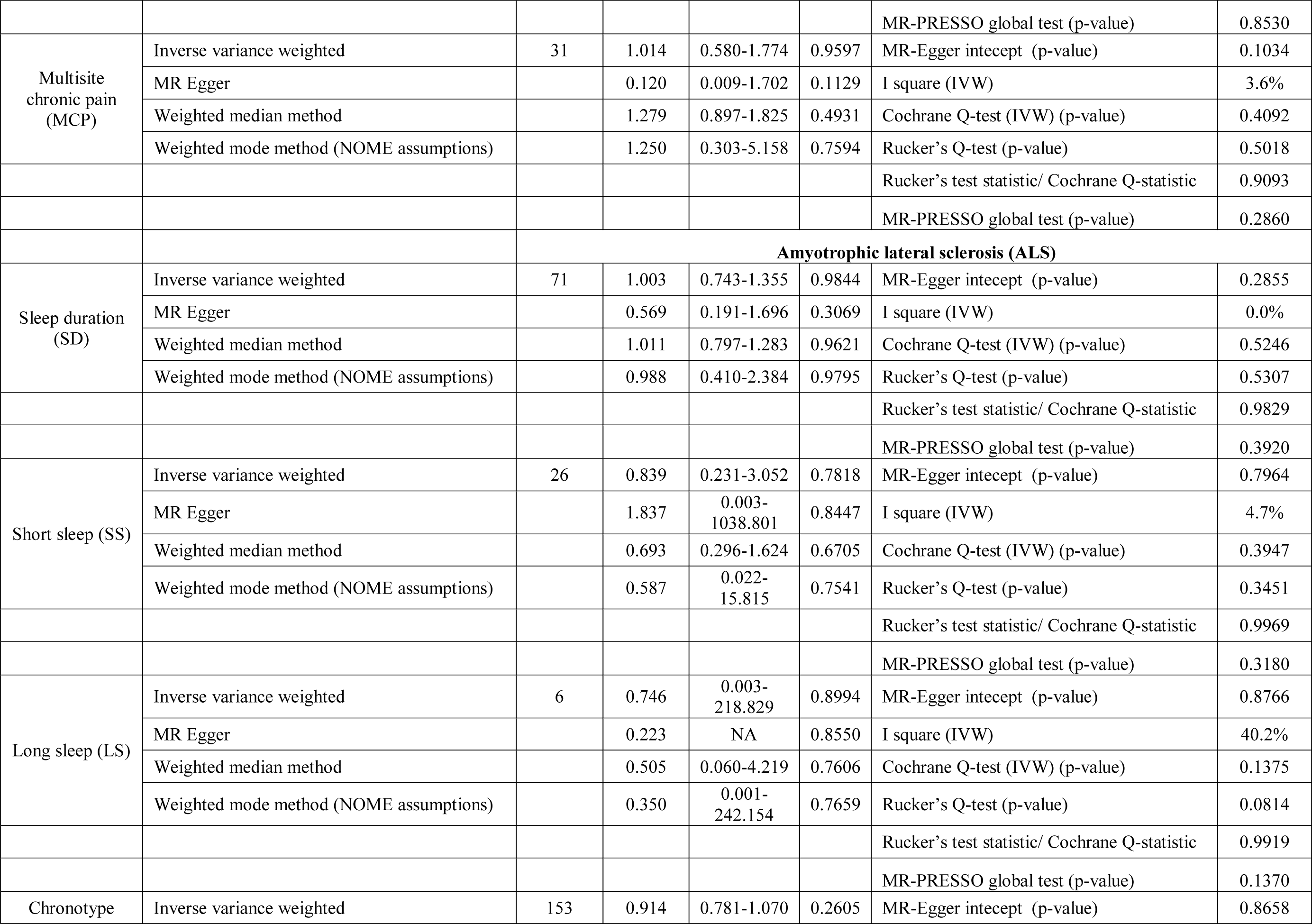
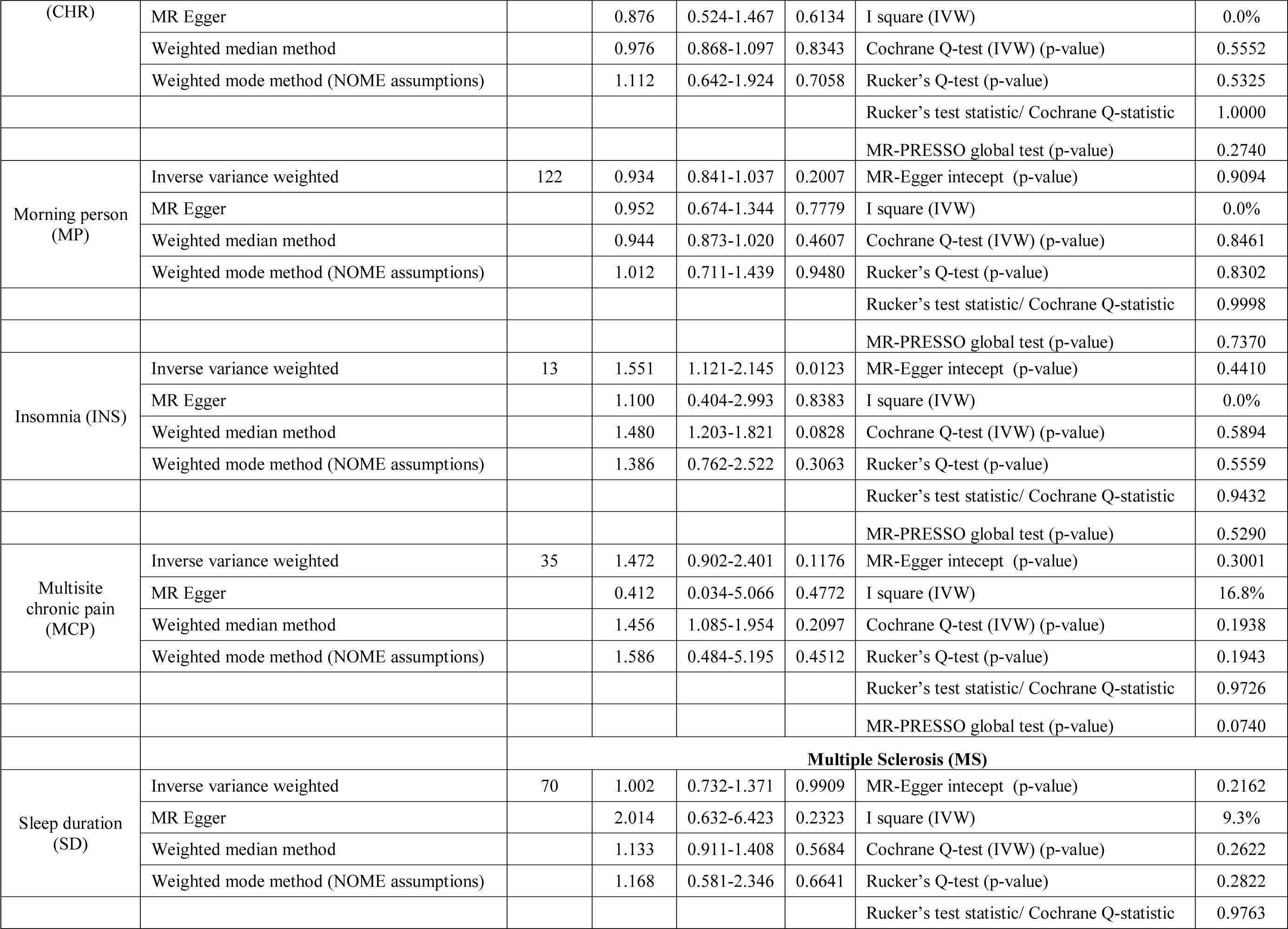
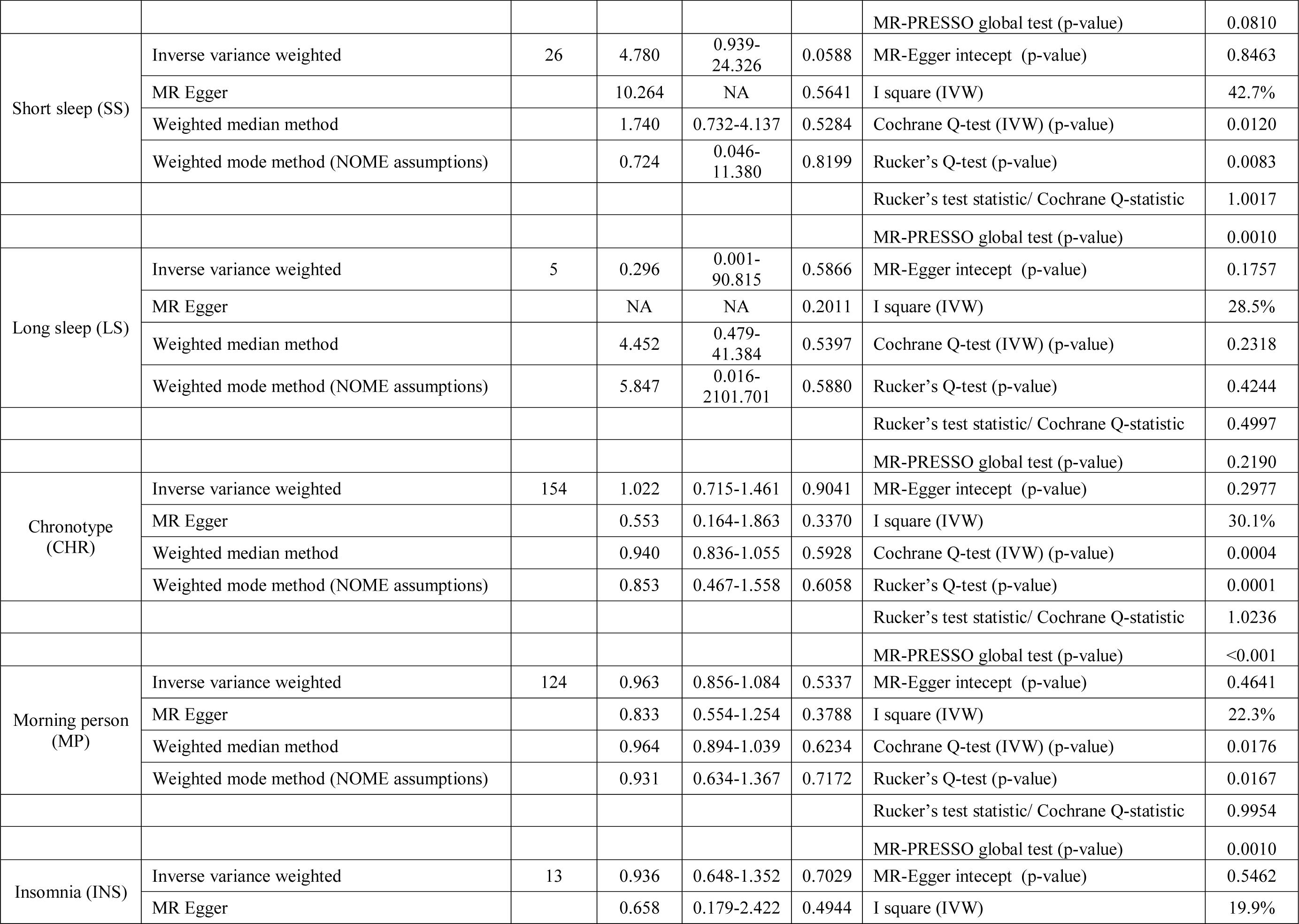
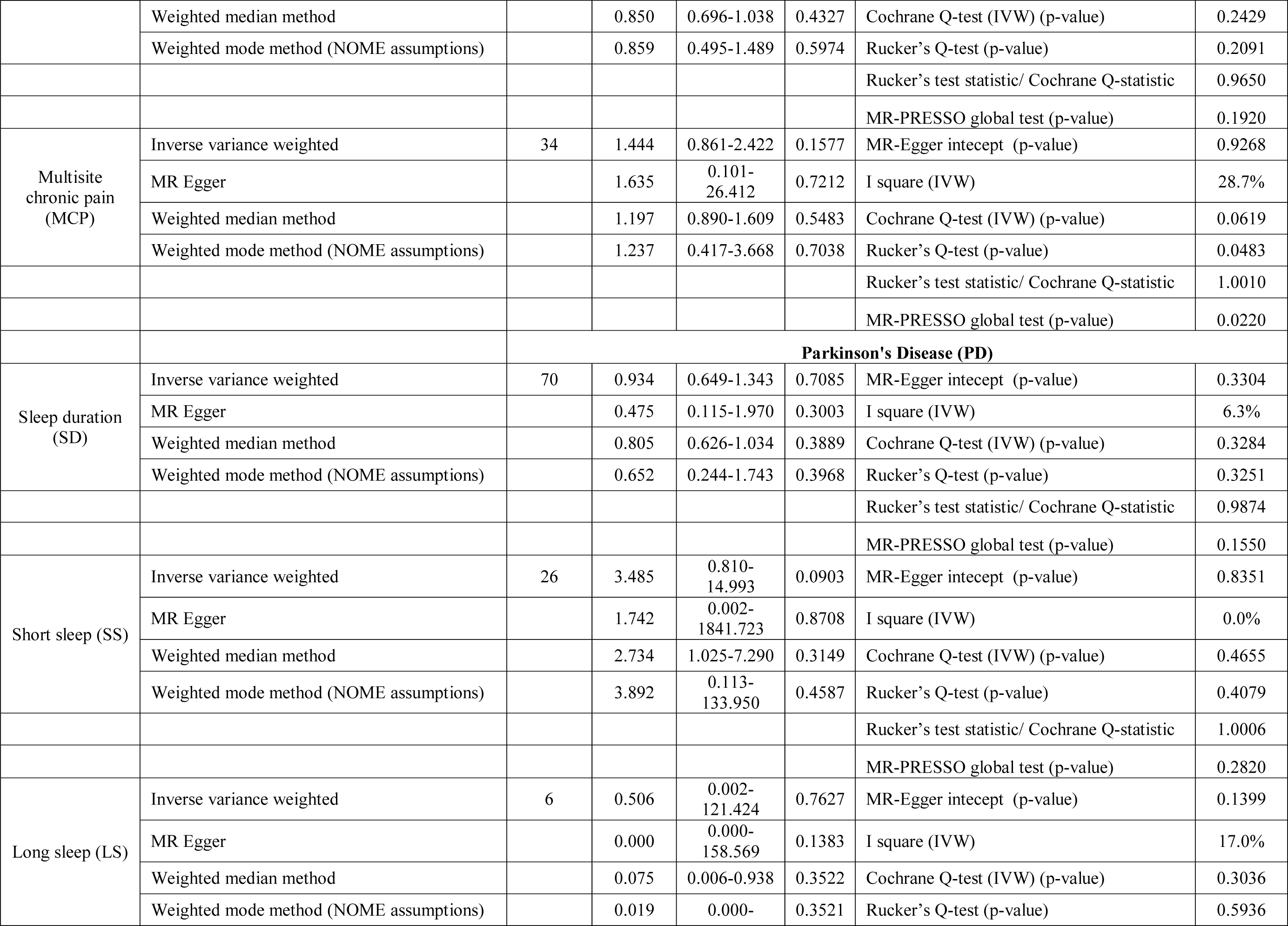
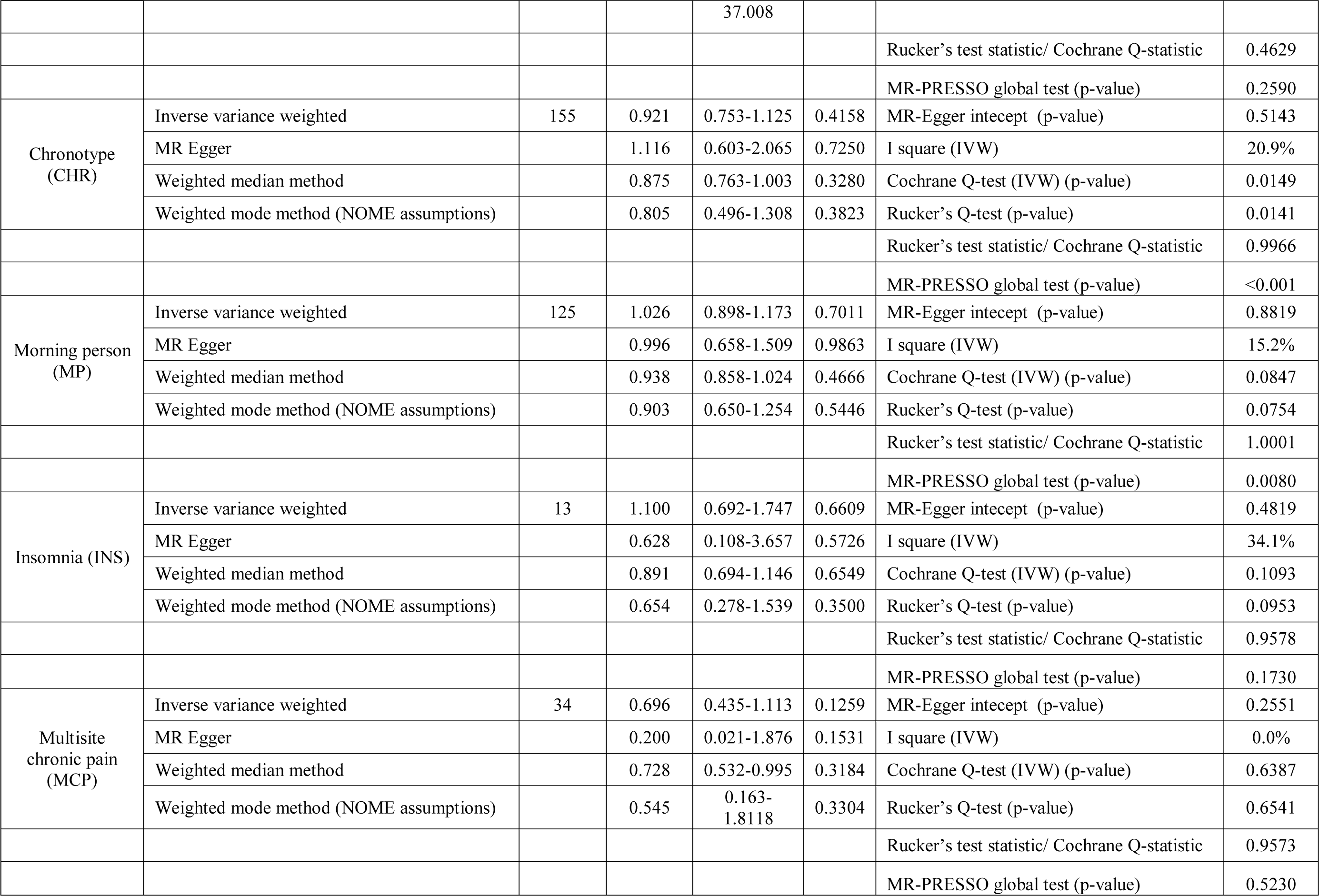
Causal effect estimates using different Mendelian randomization (MR) methods and heterogeneity analysis of causal effect estimates for neurodegenerative disorders (NDDs) using various sleep and pain-related traits as exposures.

#### AMD

We observed a highly significant causal effect of MP on AMD with morning persons (OR_IVW_ = 1.19, 95% CI 1.08, 1.32, P = 0.00073). Heterogeneity check confirmed the reliability of the observed association with absence of any heterogeneity in the distribution of effect estimates of individual genetic variants (I^2^ = 0.0%, Cochrane P = 0.929, Rucker’s Q-test P = 0.9414, MR-PRESSO global test P = 0.8420). The distribution of individual SNP-level effect estimates and the effect estimates computed through different MR methods for the effect of MP on AMD is further shown as scatter and funnel plots in **Figure 1**. We observed a similar directionality of causal effect estimates using the WME method (OR_WME_ = 1.126, 95% CI = 1.04,1.21). We also observed a similar trend using a highly correlated but continuous trait CHR on AMD (OR_IVW_ = 1.27, 95% CI 1.08, 1.49, P = 0.0034). The directionality of findings was further confirmed by significantly higher variance explained by genetic instruments for MP and CHR than that explained by the respective genetic instruments for AMD (P_Steiger_=2.1x10^-98^ and P_Steiger_=1.65x10^- 24^). In contrast, we did not observe any direct role of pain on predisposition to AMD.

**Figure 1.**
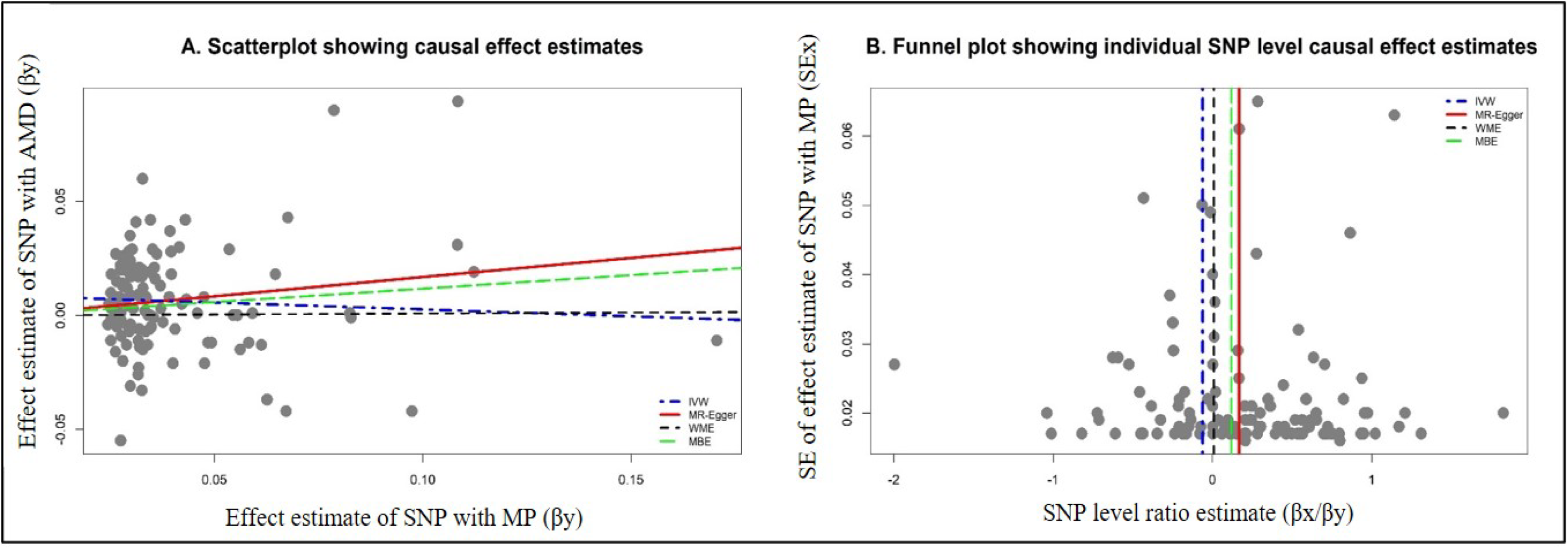
Graphical representation of causal association analysis and assessment of pleiotropy **A.** Scatter plot showing causal effect estimates computed using various MR methods for the association of morning person (MP) as exposure with age-related macular degeneration (AMD) as an outcome. **B.** Funnel plot showing the extent of heterogeneity among the individual Wald ratio estimates for the morning person (MP) as exposure with age-related macular degeneration (AMD) as an outcome. *IVW: Inverse variance weighted method, WME: Weighted median method, MBE: Weighted mode method (NOME assumptions)*

#### AD

We observed a suggestive risky causal effect of SS on AD (OR_IVW_ = 1.26, 95% CI 1.08, 1.46, P = 0.0044). Heterogeneity check further confirmed the reliability of the observed association with absence of any heterogeneity in the distribution of effect estimates of individual genetic variants (I^2^ = 0.0%, Cochrane P = 0.595, Rucker’s Q-test P = 0.5279, MR-PRESSO global test P = 0.4520). A similar directionality in causal effect estimates was also observed using the WME method (OR=1.12, 95% CI 1.10, 1.35). However, we did not observe any role of pain on predisposition to AD.

#### ALS

We observed a suggestive risky causal effect of INS on ALS (OR_IVW_ = 1.55, 95% CI 1.12, 2.14, P = 0.0123). On the other hand, we failed to observe any role of pain on predisposition to ALS.

#### MS

We did not observe any direct role of sleep and pain-related traits on predisposition to MS.

#### PD

Similar to the absence of the effect of sleep and pain-related traits on MS, our MR analysis failed to detect the role of sleep and pain-related traits on predisposition to PD.

### Sensitivity analysis

#### Exclusion of overlapping samples

Concerning direct MR, the association of SS with AD was lost after the exclusion of overlapping UKB samples (data not shown). In the reverse MR, PD showed a suggestion of a strong protective effect against CHR and MP after the exclusion of overlapping UKB samples (data not shown).

#### Reverse causality check

Reverse causal estimates for various sleep and pain-related traits using various NDDs as exposure have been shown in **Table 3**.

**Table 3.**
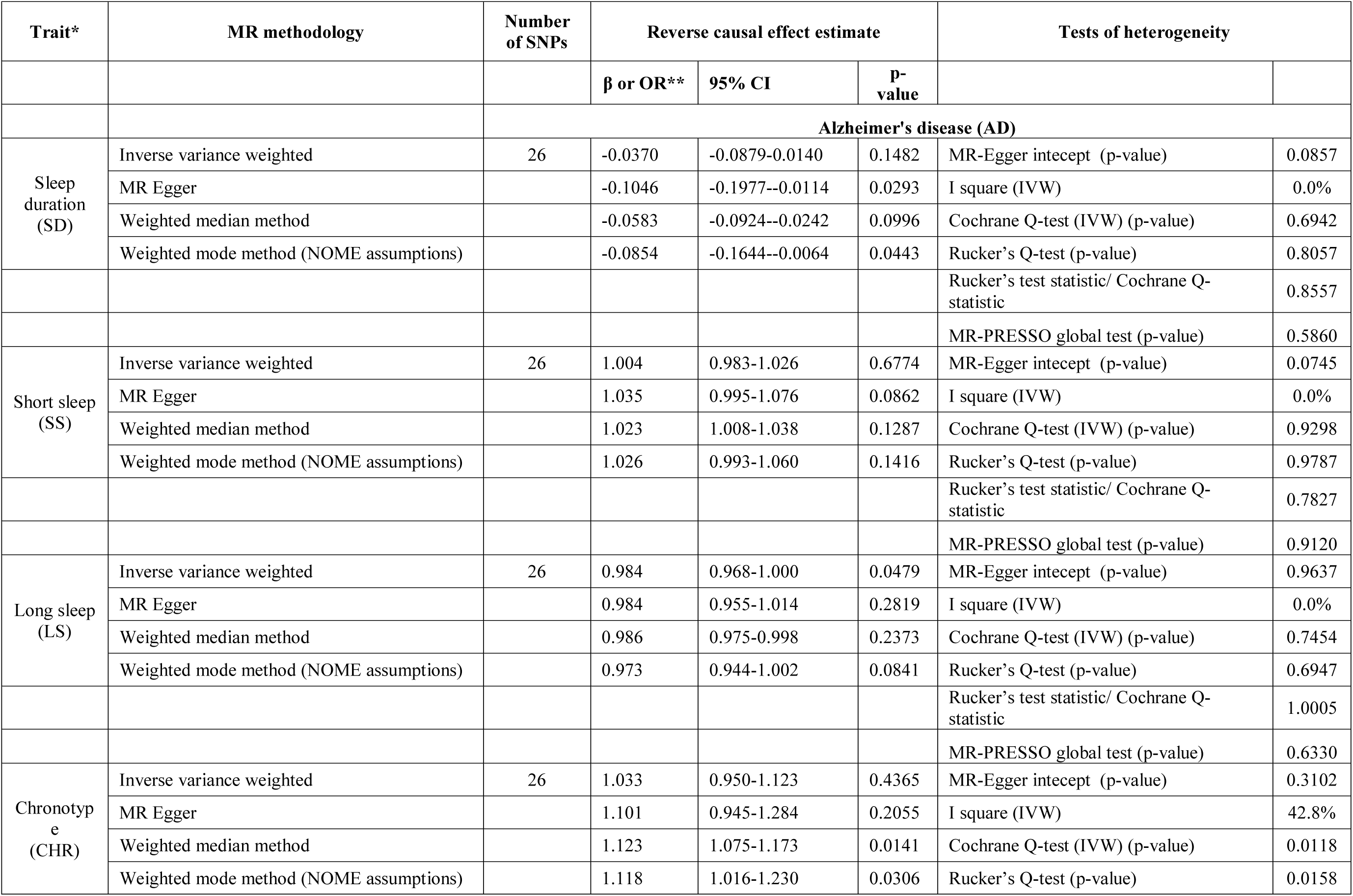
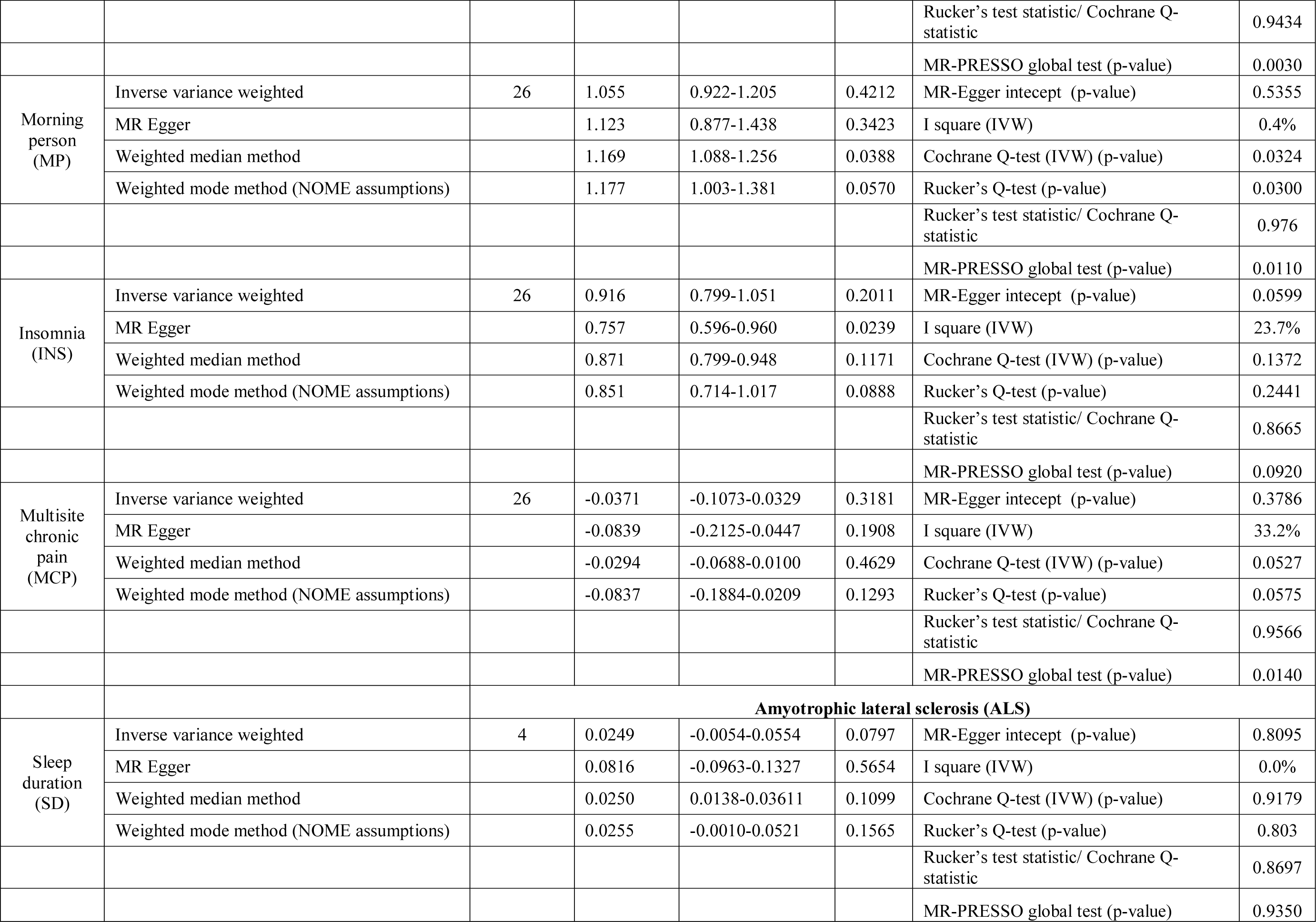
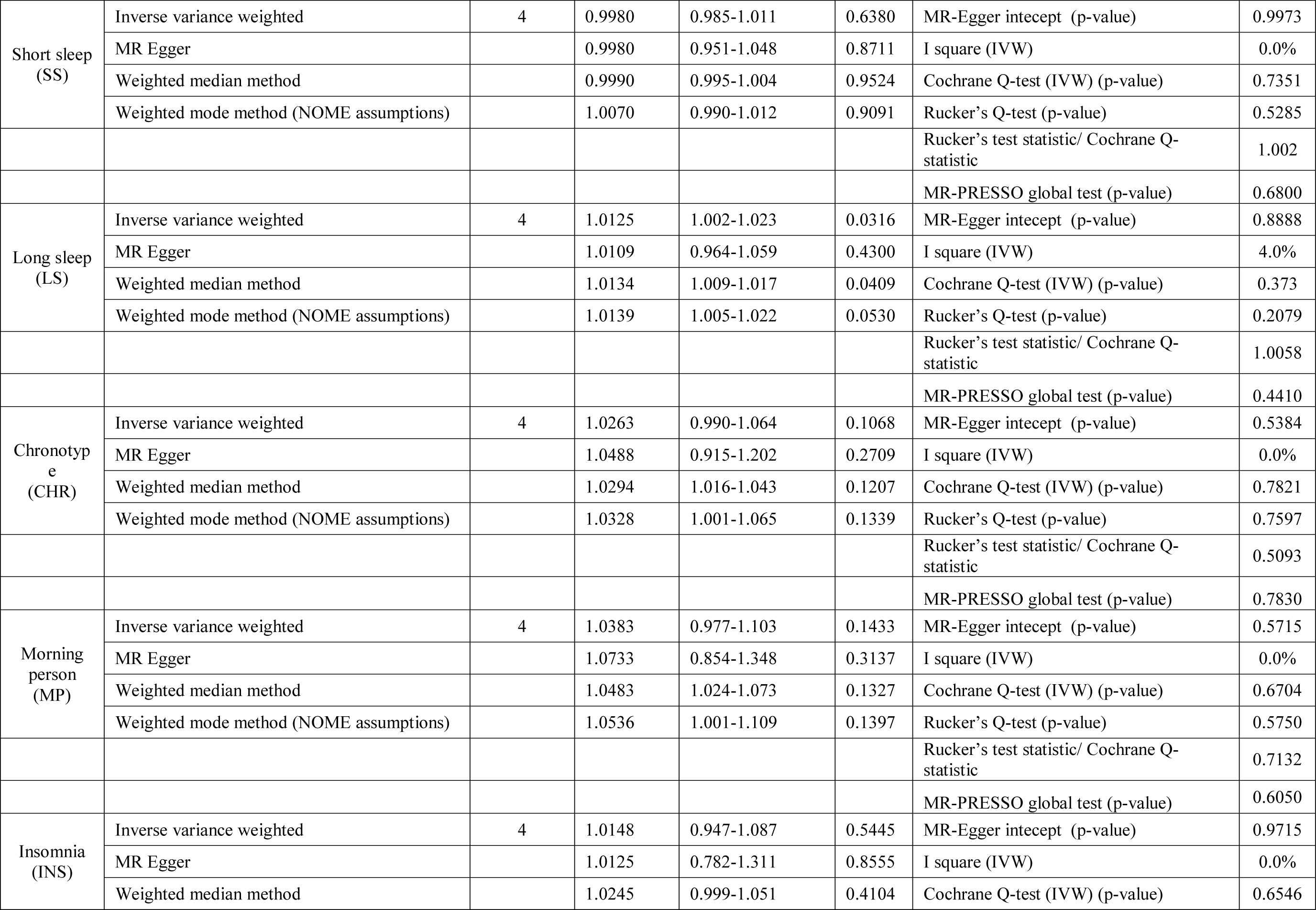
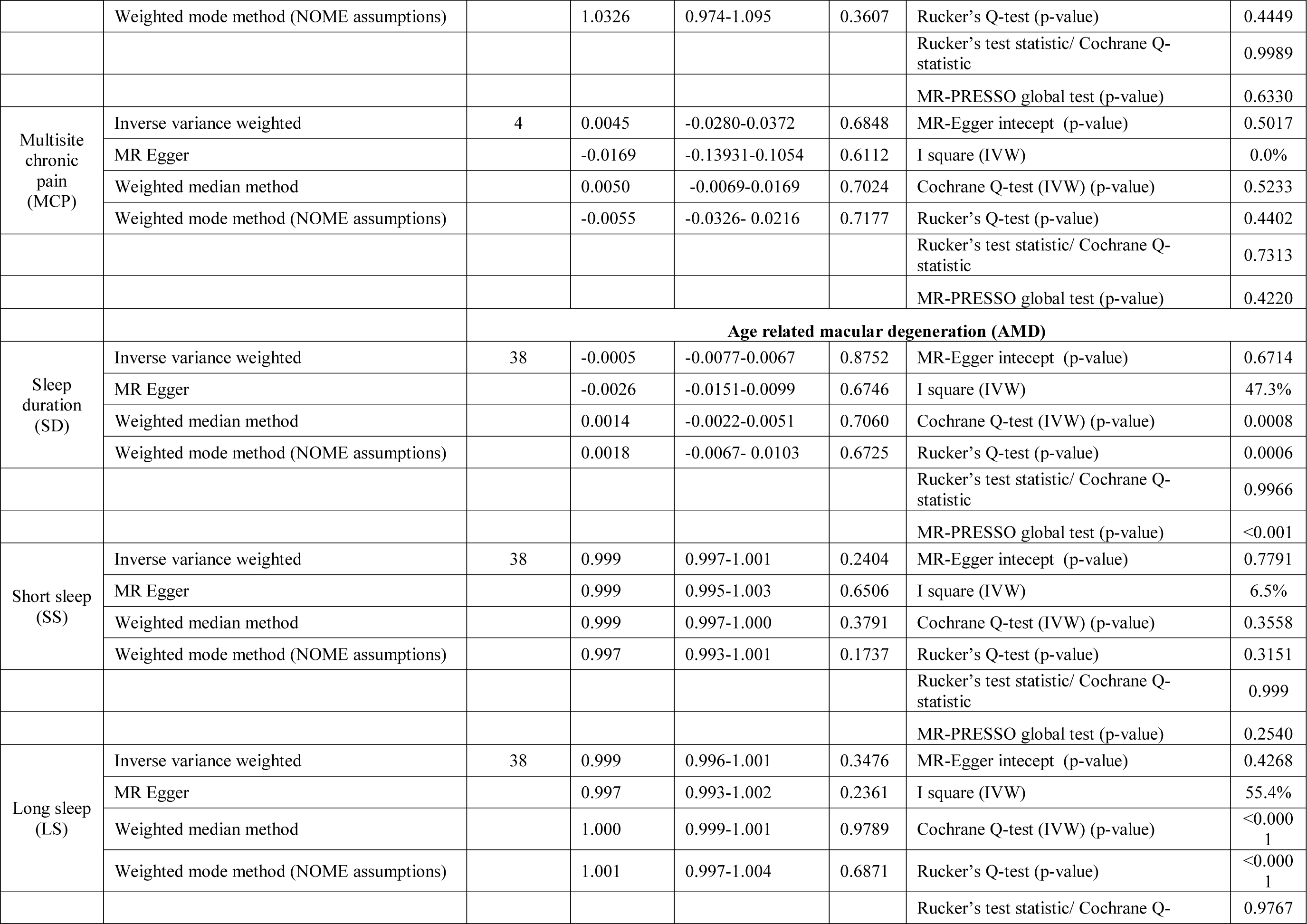
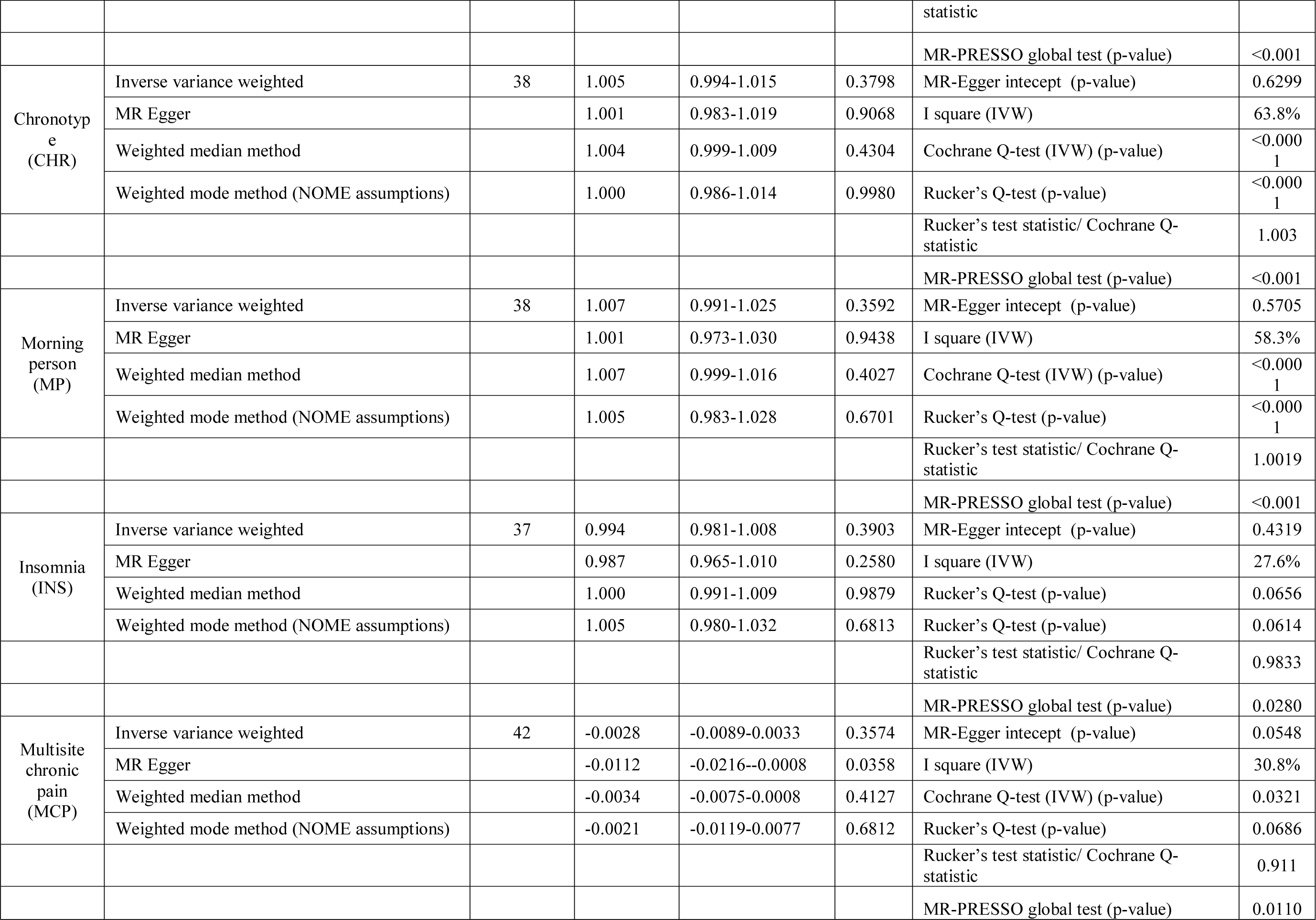
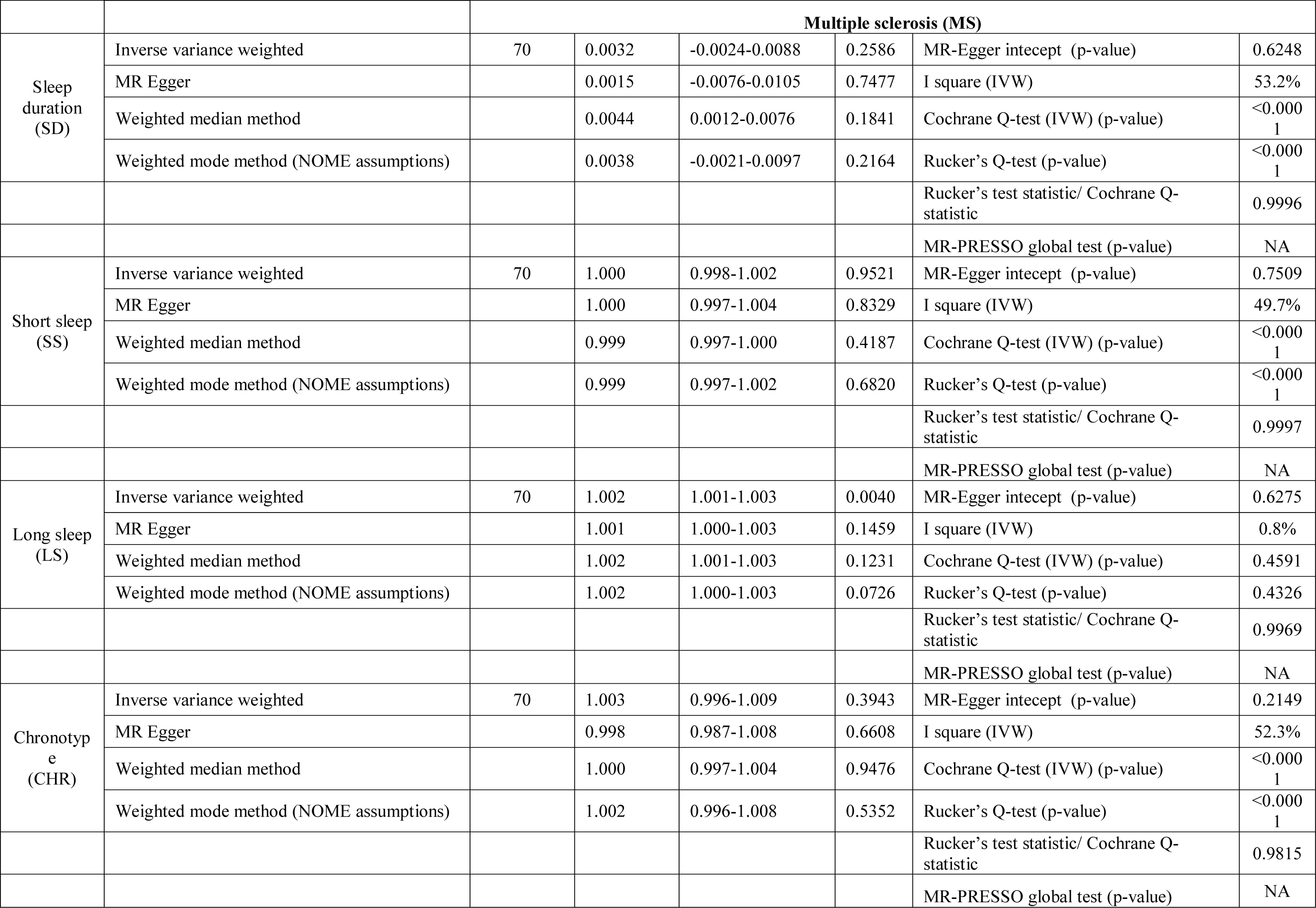
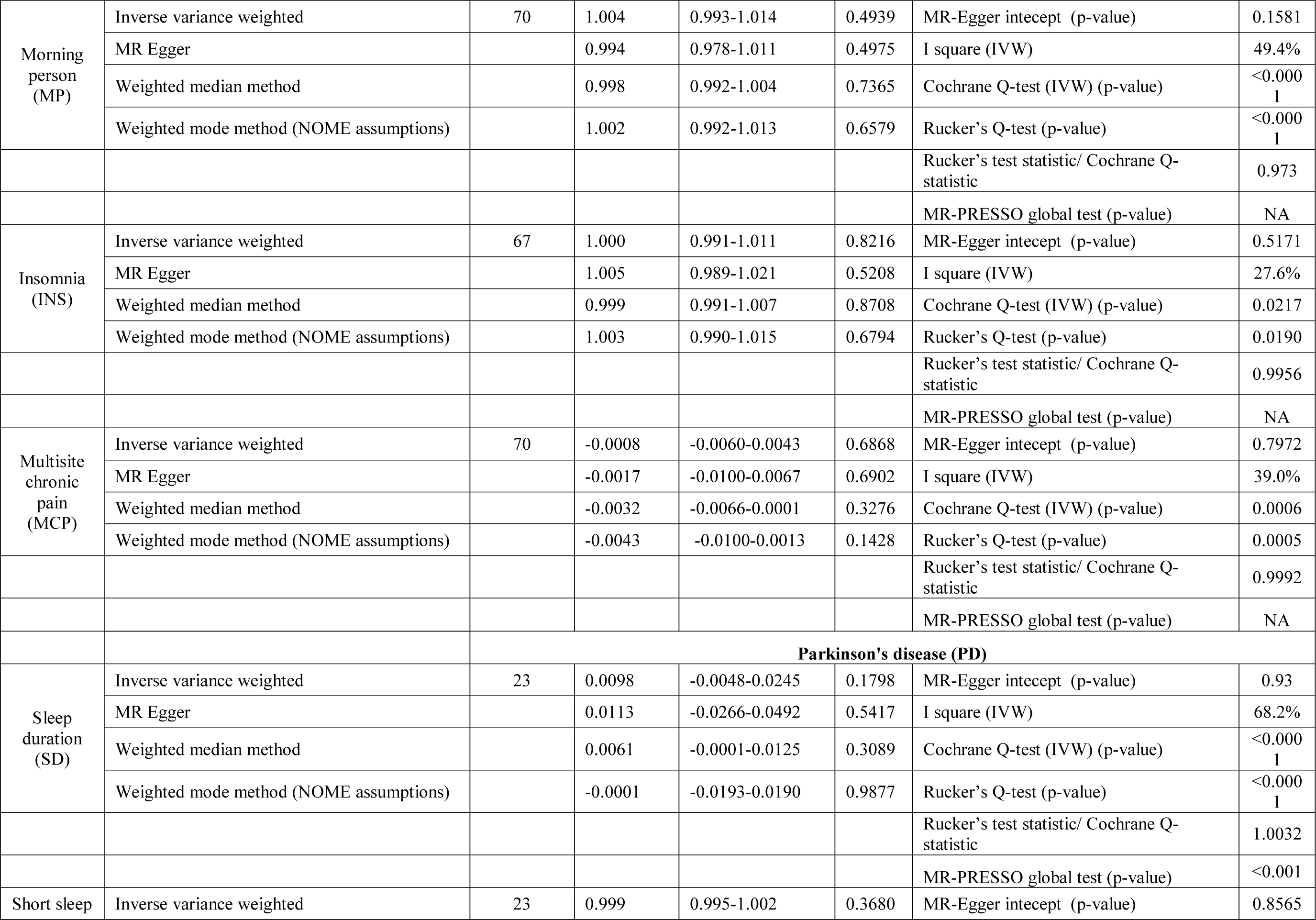
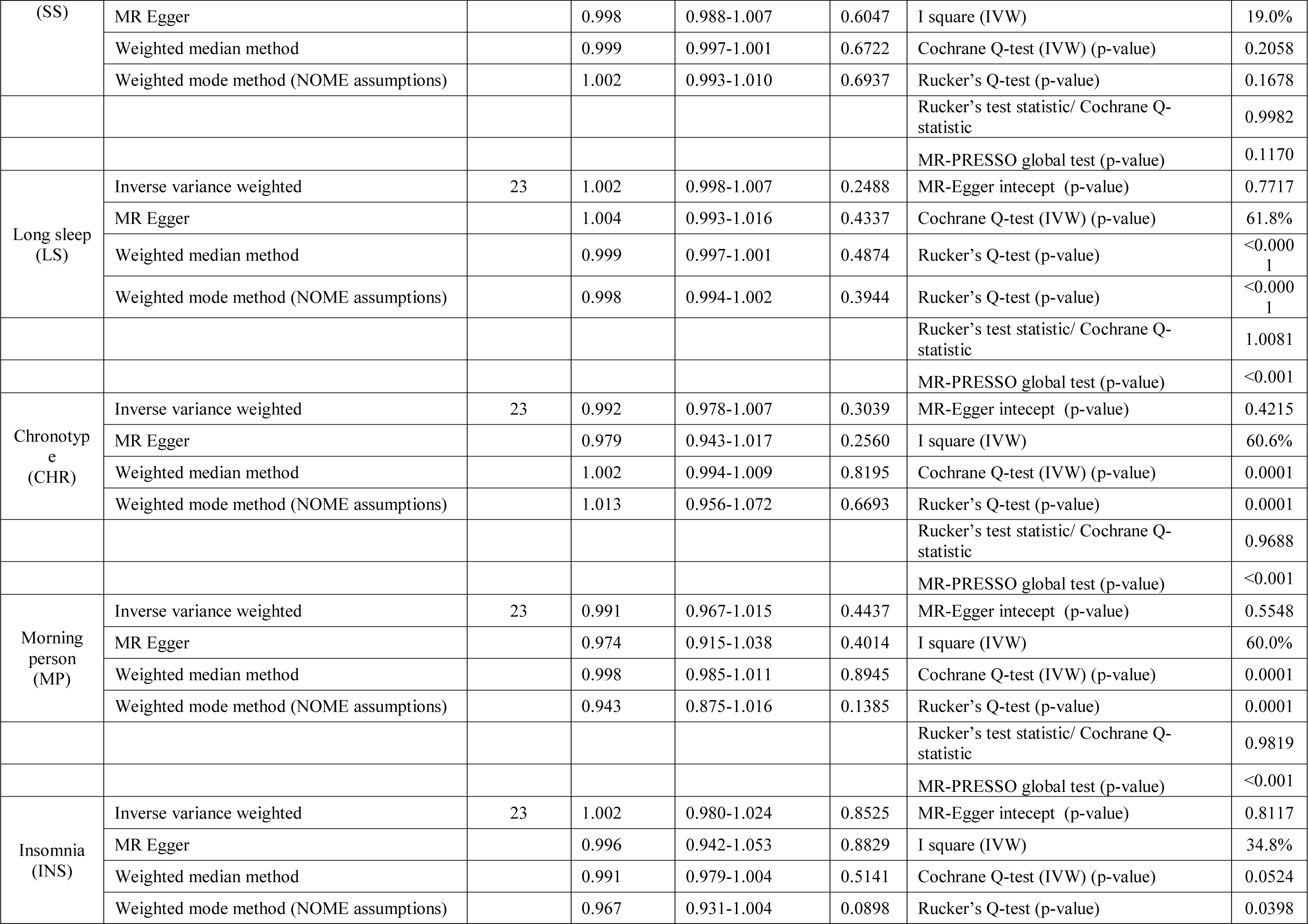
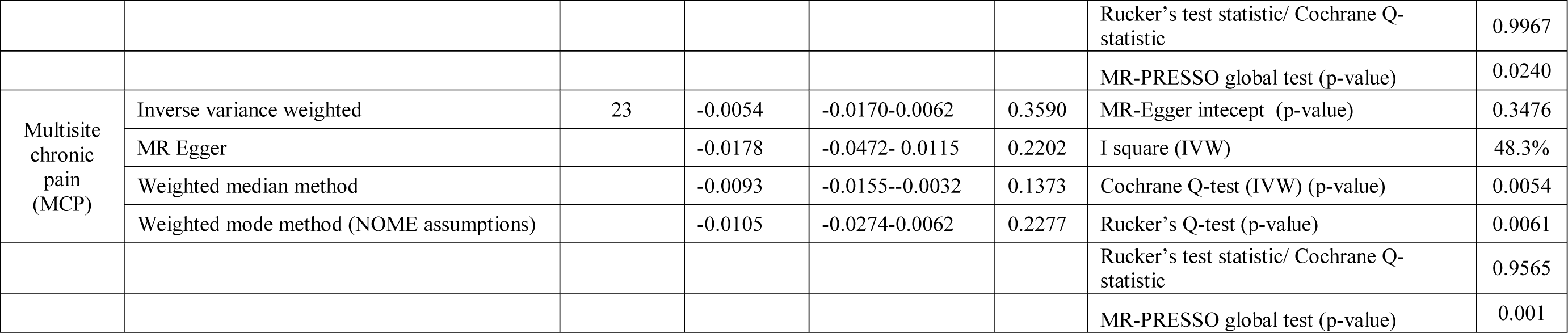
Causal effect estimates using different Mendelian randomization (MR) methods and heterogeneity analysis of causal effect estimates for various sleep and pain-related traits using Neurodegenerative disorders (NDDs) as exposures.

#### AMD

Our reverse casual check confirmed the directionality of the observed associations of MP and CHR with AMD as we failed to observe any effect of AMD on MP and CHR.

#### AD

Our reverse causal check confirmed the role of SS in predisposition to AD as we failed to observe the causal effect of AD on SS. Interestingly, all the sleep-related traits except SS were observed to be influenced by a genetic predisposition to AD when employing non-IVW methods for judging causal effects of sleep-related traits on AD.

#### ALS

Our reverse casual check confirmed the role of INS on predisposition to ALS. On the contrary, our findings suggested a causal role of genetic predisposition to ALS on LS with a consistent significant risk effect using IVW, WME, and MBE methods.

#### Exclusion of potential pleiotropic variants

##### Leave-one-out-approach

We failed to observe the predominant influence of any of the variants on causal effect estimates of MP with AMD, as shown in **Table 4**. Similarly, the observed associations of CHR with AMD, SS with AD, and INS with ALS were retained (**Supplementary Table 3**).

**Table 4.** Sensitivity analysis of the causal effect of a morning person (MP) trait on predisposition to age- related macular degeneration (AMD) using leave-one-out SNP method

| Excluded SNP | IVW OR | 95% CI | p-value |
| --- | --- | --- | --- |
| rs17374439 | 1.196 | 1.081-1.323 | 0.0007 |
| rs3767240 | 1.180 | 1.067-1.305 | 0.0015 |
| rs12140153 | 1.195 | 1.080-1.321 | 0.0006 |
| rs7547493 | 1.200 | 1.085-1.328 | 0.0005 |
| rs11588913 | 1.191 | 1.077-1.317 | 0.0008 |
| rs72720396 | 1.181 | 1.067-1.306 | 0.0015 |
| rs4949980 | 1.193 | 1.079-1.319 | 0.0007 |
| rs17575798 | 1.192 | 1.077-1.318 | 0.0008 |
| rs6537834 | 1.194 | 1.079-1.320 | 0.0007 |
| rs11587758 | 1.187 | 1.073-1.313 | 0.0010 |
| rs75650221 | 1.191 | 1.077-1.317 | 0.0008 |
| rs11580135 | 1.188 | 1.074-1.314 | 0.0009 |
| rs1144566 | 1.201 | 1.085-1.330 | 0.0005 |
| rs10196909 | 1.199 | 1.084-1.326 | 0.0005 |
| rs75120545 | 1.199 | 1.084-1.326 | 0.0005 |
| rs786406 | 1.191 | 1.077-1.317 | 0.0008 |
| rs10495976 | 1.190 | 1.076-1.316 | 0.0009 |
| rs7602425 | 1.189 | 1.075-1.315 | 0.0009 |
| rs778147 | 1.196 | 1.081-1.323 | 0.0006 |
| rs6744983 | 1.188 | 1.075-1.314 | 0.0009 |
| rs10520176 | 1.193 | 1.079-1.320 | 0.0007 |
| rs28380327 | 1.198 | 1.083-1.325 | 0.0006 |
| rs1947198 | 1.188 | 1.075-1.314 | 0.0009 |
| rs11679484 | 1.184 | 1.070-1.309 | 0.0012 |
| rs138964083 | 1.193 | 1.079-1.320 | 0.0007 |
| rs77008212 | 1.196 | 1.080-1.323 | 0.0007 |
| rs114870822 | 1.191 | 1.077-1.317 | 0.0008 |
| rs34581681 | 1.194 | 1.080-1.321 | 0.0007 |
| rs13059636 | 1.203 | 1.087-1.330 | 0.0004 |
| rs115774037 | 1.187 | 1.074-1.313 | 0.0010 |
| rs11712056 | 1.196 | 1.081-1.323 | 0.0006 |
| rs60194061 | 1.189 | 1.075-1.315 | 0.0009 |
| rs957501 | 1.198 | 1.083-1.325 | 0.0006 |
| rs13065394 | 1.188 | 1.075-1.314 | 0.0009 |
| rs2102506 | 1.195 | 1.081-1.322 | 0.0006 |
| rs3850174 | 1.199 | 1.084-1.326 | 0.0005 |
| rs2239626 | 1.196 | 1.081-1.323 | 0.0006 |
| rs34627176 | 1.200 | 1.085-1.327 | 0.0005 |
| rs231398 | 1.194 | 1.079-1.320 | 0.0007 |
| rs12498561 | 1.192 | 1.078-1.318 | 0.0008 |
| rs17292170 | 1.191 | 1.077-1.317 | 0.0008 |
| rs7691121 | 1.197 | 1.082-1.323 | 0.0006 |
| rs4339281 | 1.186 | 1.073-1.312 | 0.0011 |
| rs4241964 | 1.206 | 1.090-1.333 | 0.0004 |
| rs10067113 | 1.192 | 1.078-1.318 | 0.0008 |
| rs7701529 | 1.194 | 1.080-1.321 | 0.0007 |
| rs34875688 | 1.188 | 1.074-1.314 | 0.0010 |
| rs1027742 | 1.189 | 1.075-1.314 | 0.0009 |
| rs2910032 | 1.192 | 1.077-1.318 | 0.0008 |
| rs9369915 | 1.193 | 1.079-1.320 | 0.0007 |
| rs2842638 | 1.188 | 1.075-1.314 | 0.0009 |
| rs2653343 | 1.202 | 1.086-1.331 | 0.0005 |
| rs60616179 | 1.195 | 1.081-1.322 | 0.0006 |
| rs520954 | 1.193 | 1.078-1.320 | 0.0008 |
| rs3877930 | 1.192 | 1.078-1.318 | 0.0008 |
| rs9365769 | 1.192 | 1.078-1.318 | 0.0008 |
| rs1494185 | 1.192 | 1.078-1.319 | 0.0008 |
| rs12669911 | 1.185 | 1.071-1.310 | 0.0011 |
| rs6967481 | 1.185 | 1.072-1.311 | 0.0011 |
| rs12704886 | 1.196 | 1.082-1.323 | 0.0006 |
| rs202157 | 1.188 | 1.074-1.314 | 0.0010 |
| rs1524472 | 1.192 | 1.078-1.319 | 0.0007 |
| rs13269289 | 1.193 | 1.079-1.319 | 0.0007 |
| rs11786306 | 1.190 | 1.076-1.316 | 0.0009 |
| rs13255030 | 1.193 | 1.079-1.320 | 0.0007 |
| rs7001604 | 1.191 | 1.077-1.317 | 0.0008 |
| rs2609589 | 1.190 | 1.076-1.316 | 0.0008 |
| rs769066 | 1.191 | 1.077-1.317 | 0.0008 |
| rs4321976 | 1.178 | 1.065-1.303 | 0.0016 |
| rs1470764 | 1.187 | 1.074-1.313 | 0.0010 |
| rs12682033 | 1.195 | 1.080-1.322 | 0.0007 |
| rs10976942 | 1.196 | 1.081-1.322 | 0.0006 |
| rs308521 | 1.188 | 1.074-1.313 | 0.0010 |
| rs62553781 | 1.194 | 1.079-1.320 | 0.0007 |
| rs10818834 | 1.188 | 1.074-1.314 | 0.0010 |
| rs34619169 | 1.192 | 1.078-1.319 | 0.0007 |
| rs28458909 | 1.184 | 1.070-1.310 | 0.0013 |
| rs2893787 | 1.192 | 1.078-1.318 | 0.0008 |
| rs4752593 | 1.190 | 1.076-1.316 | 0.0009 |
| rs6599694 | 1.195 | 1.080-1.321 | 0.0007 |
| rs72632979 | 1.193 | 1.079-1.319 | 0.0007 |
| rs10501087 | 1.182 | 1.069-1.308 | 0.0013 |
| rs512647 | 1.194 | 1.080-1.321 | 0.0007 |
| rs11032362 | 1.206 | 1.090-1.335 | 0.0004 |
| rs11229543 | 1.191 | 1.077-1.318 | 0.0008 |
| rs4936291 | 1.194 | 1.080-1.320 | 0.0007 |
| rs73606718 | 1.197 | 1.082-1.323 | 0.0006 |
| rs2467109 | 1.195 | 1.081-1.322 | 0.0006 |
| rs7302062 | 1.188 | 1.074-1.314 | 0.0010 |
| rs7316768 | 1.199 | 1.084-1.326 | 0.0005 |
| rs7313852 | 1.207 | 1.091-1.336 | 0.0004 |
| rs11174781 | 1.192 | 1.078-1.319 | 0.0008 |
| rs7959983 | 1.198 | 1.083-1.325 | 0.0006 |
| rs7304278 | 1.188 | 1.074-1.313 | 0.0010 |
| rs9597241 | 1.191 | 1.077-1.318 | 0.0008 |
| rs9573971 | 1.180 | 1.067-1.306 | 0.0015 |
| rs11841335 | 1.192 | 1.077-1.318 | 0.0008 |
| rs10149448 | 1.191 | 1.077-1.317 | 0.0008 |
| rs12927162 | 1.194 | 1.079-1.321 | 0.0007 |
| rs1421085 | 1.182 | 1.068-1.308 | 0.0014 |
| rs62046253 | 1.188 | 1.074-1.314 | 0.0009 |
| rs11645898 | 1.189 | 1.075-1.315 | 0.0009 |
| rs2518022 | 1.204 | 1.088-1.332 | 0.0004 |
| rs3760185 | 1.190 | 1.076-1.317 | 0.0008 |
| rs72829936 | 1.187 | 1.073-1.313 | 0.0010 |
| rs35653190 | 1.186 | 1.073-1.312 | 0.0010 |
| rs17682747 | 1.193 | 1.078-1.319 | 0.0007 |
| rs2949923 | 1.189 | 1.075-1.315 | 0.0009 |
| rs62082401 | 1.192 | 1.078-1.318 | 0.0008 |
| rs1013987 | 1.193 | 1.078-1.319 | 0.0007 |
| rs4419127 | 1.194 | 1.079-1.321 | 0.0007 |
| rs12969848 | 1.184 | 1.070-1.310 | 0.0012 |
| rs9962650 | 1.186 | 1.072-1.311 | 0.0011 |
| rs9964420 | 1.197 | 1.082-1.324 | 0.0006 |
| rs11152350 | 1.189 | 1.075-1.315 | 0.0009 |
| rs9636202 | 1.198 | 1.083-1.325 | 0.0005 |
| rs11670534 | 1.196 | 1.082-1.323 | 0.0006 |
| rs78095690 | 1.187 | 1.073-1.313 | 0.0010 |
| rs2072727 | 1.186 | 1.072-1.311 | 0.0011 |
| rs139911 | 1.196 | 1.081-1.323 | 0.0006 |
| rs4822107 | 1.187 | 1.074-1.313 | 0.0010 |

##### Leave-one-group out-approach

The cumulative influence of potential pleiotropic SNPs identified through a comprehensive screening of the Phenoscanner database on observed associations is summarized in **Table 5**. We identified 46 and 51 SNPs used for causal effect estimates of MP and CHR with AMD, respectively, as an outcome as potential pleiotropic SNPs. However, the exclusion of these SNPs did not influence the observed casual association of MP and CHR with AMD (OR = 1.202, 95% CI 1.0549, 1.370; OR = 1.262, 95% CI 1.049, 1.520). On the contrary, associations of SS with AD and INS with ALS were lost, which could be attributed to the presence of a high proportion of pleiotropic SNPs in the genetic instruments for SS and INS.

**Table 5.** Sensitivity analysis of causal effect estimates of sleep-related traits on neurodegeneration using leave-group-out SNP method by excluding potential pleiotropic SNPs identified through Phenoscanner.

| Exposure | Outcome | Number of SNPs in the genetic instrument | Number of pleiotropic SNPs (Phenoscanner) | Number of SNPs remaining | OR | 95% CI | p-value |
| --- | --- | --- | --- | --- | --- | --- | --- |
| Morning person (MP) | Age related macular degeneration (AMD) | 121 | 46 | 75 | 1.202 | 1.055-1.370 | 0.0063 |
| Chronotype (CHR) | Age related macular degeneration (AMD) | 150 | 51 | 99 | 1.262 | 1.049-1.520 | 0.0141 |
| Short sleep (SS) | Alzheimer's disease (AD) | 26 | 14 | 12 | 1.041 | 0.819-1.320 | 0.723 |
| Insomnia (INS) | Amyotrophic lateral sclerosis (ALS) | 13 | 10 | 3 | 2.135 | 0.457-9.979 | 0.168 |

#### Influence of regional brain expression

The cumulative influence of brain-region-specific quantitative trait variants retrieved from the Genotype-Tissue Expression Project is summarized in **Table 6**. We specifically identified a high proportion of SNPs influencing brain expression in the cerebellum and basal ganglia region. However, the exclusion of these SNPs did not affect the overall causal association of CHR and MP with AMD. Similarly, we failed to observe the effect of any of the other brain regions on the observed associations. We also failed to observe any influence of brain region specific expression on other observed associations (data not shown).

**Table 6.** Sensitivity analysis of causal effect estimates of sleep-related traits on neurodegeneration by exploring the potential influence of specific brain regions using variants involved in the regional expression.

| Brain region | Causal effect estimates of MP with AMD |  |  |  |  | Causal effect estimates of CHR with AMD |  |  |  |  |
| --- | --- | --- | --- | --- | --- | --- | --- | --- | --- | --- |
|  | Number of SNPs involved in expression | Number of SNPs remaining | IVW OR | 95% CI | p-value | Number of SNPs involved in expression | Number of SNPs | IVW OR | 95% CI | p-value |
| Amygdala | 5 | 116 | 1.184 | 1.069-1.312 | 0.0014 | 6 | 144 | 1.245 | 1.061-1.462 | 0.0077 |
| Anterior cingulate cortex (BA24) | 8 | 113 | 1.188 | 1.070-1.318 | 0.0014 | 12 | 138 | 1.269 | 1.088-1.479 | 0.0027 |
| Brain - Caudate (basal ganglia) | 14 | 107 | 1.180 | 1.061-1.313 | 0.0027 | 19 | 131 | 1.262 | 1.075-1.482 | 0.0049 |
| Brain - Cerebellar Hemisphere | 13 | 108 | 1.185 | 1.066-1.317 | 0.0019 | 17 | 133 | 1.285 | 1.098-1.504 | 0.0020 |
| Brain - Cerebellum | 16 | 105 | 1.186 | 1.065-1.320 | 0.0021 | 21 | 129 | 1.271 | 1.079-1.497 | 0.0044 |
| Brain - Cortex | 13 | 108 | 1.175 | 1.058-1.306 | 0.0030 | 17 | 133 | 1.252 | 1.071-1.462 | 0.0050 |
| Brain - Cerebellar Hemisphere | 13 | 108 | 1.185 | 1.066-1.317 | 0.0019 | 17 | 133 | 1.285 | 1.098-1.504 | 0.0020 |
| Brain - Frontal Cortex (BA9) | 14 | 107 | 1.176 | 1.057-1.307 | 0.0031 | 15 | 135 | 1.264 | 1.083-1.476 | 0.0033 |
| Brain - Hippocampus | 7 | 114 | 1.197 | 1.080-1.328 | 0.0008 | 11 | 139 | 1.296 | 1.111-1.511 | 0.0011 |
| Brain - Hypothalamus Brain | 0 | 121 | 1.192 | 1.078-1.318 | 0.0007 | 0 | 150 | 1.269 | 1.083-1.486 | 0.0034 |
| Brain - Nucleus accumbens (basal ganglia) | 12 | 109 | 1.189 | 1.070-1.320 | 0.0015 | 17 | 133 | 1.268 | 1.082-1.486 | 0.0037 |
| Brain - Putamen (basal ganglia) | 8 | 113 | 1.195 | 1.078-1.326 | 0.0009 | 12 | 138 | 1.266 | 1.086-1.475 | 0.0028 |
| Brain - Spinal cord (cervical c-1) | 5 | 116 | 1.198 | 1.081-1.326 | 0.0007 | 9 | 141 | 1.295 | 1.113-1.508 | 0.0010 |
| Brain - Substantia nigra | 2 | 119 | 1.200 | 1.084-1.328 | 0.0005 | 5 | 145 | 1.301 | 1.121-1.511 | 0.0007 |

#### Discussion

The use of GWAS data in MR-based approaches has opened up opportunities to assess and define clinically relevant signatures for a diverse spectrum of diseases. Our study supports the role of a person’s underlying circadian rhythm in genetic predisposition to neurodegeneration. We found an association of genetically predicted morning person trait with AMD. The correlated trait CHR also had a suggestive risky association with AMD. We also found suggestive evidence for a possible association of genetically predicted SS with AD and INS with ALS. Surprisingly, our study, however, found no evidence to support the association between pain and NDD.

### Comparison with other studies

To date, evidence from observational studies has shown a remarkable heterogeneity in the association of different circadian traits with various NDDs. A recent study investigating the incidence of AMD in 108,225 participants observed that INS patients were 33% more likely to have subsequent AMD (HR 1.33, 95% CI 1.18, 1.48)^5^. Previously, an observational study of 57 patients with neovascular AMD and 108 controls found a significantly increased risk of neovascular AMD in patients sleeping less than 6 hours compared to those sleeping 7-8 hours (OR 3.29, 95% CI 1.32, 8.27)^64^. Another study failed to detect an association with LS in 316 patients with neovascular AMD compared to 500 patients without AMD^65^. However, the study did find an association of LS with geographic atrophy, an advanced form of AMD in 61 individuals (presence of a discrete area of atrophy with a diameter of 175 m). A recent observational study further reported that individuals who take an afternoon nap are 60% less likely to be diagnosed with late AMD (56 late AMD vs 1204 No AMD)^66^. As darkness is known to stimulate the secretion of melatonin from the pineal gland, our findings are in agreement with previous studies showing that increased melatonin synthesis could play a protective role in the pathophysiology of AMD^67^. However, a recent randomized controlled trial (RCT) failed to show any beneficial effect of low-level night-time light therapy on the progression of AMD^68^.

In contrast to previously reported findings from the epidemiological studies, we failed to observe any association of INS, SS, and LS with AMD using the genetic data in the present study. However, we observed that MP is more likely to be predisposed to AMD (OR 1.19, 95% CI 1.08, 1.32). Our study suggests that more prolonged exposure to daylight in such individuals could increase the risk for AMD. However, our findings are in contrast to a recent meta-analysis of observational studies demonstrating an absence of association between sunlight exposure and AMD (OR 1.12 95% CI 0.76, 1.67)^69^. One of the possible reasons for this discrepancy could be that only one of the 14 studies included in the meta-analysis was a cohort study. The only included cohort study was a ten-year follow-up study which demonstrated that participants exposed to summer sun for more than 5 hours a day were more likely to show increased retinal pigment (RR 2.99, 95% CI 1.18, 7.60) and develop early age-related maculopathy (RR 2.20, 95% CI 1.02, 4.73) in comparison to those exposed to less than 2 hours per day ^70^. It has also been suggested that excessive light exposure may induce phototoxic damage to the retinal pigmental epithelium and possibly contribute to the gradual worsening of vision in AMD^71–73^.

To further substantiate our findings implicating MP on the risk of developing AMD; the use of heterogeneity checking approaches such as MR-Egger intercept test and Cochran Q-statistic, and cross-validation approaches such as leave-one-SNP-out and leave-one-group-out method, highlighting the robustness of our findings showing that MP and CHR could alter risk to AMD rather than INS or SD.

Compared to the impact of circadian rhythms in other NDDs, the role of sleep-related traits has been well investigated in AD, with mixed findings. Previous studies have predominantly focused on sleep-wake rhythmicity, showing a higher incidence of sleep fragmentations and lower amplitude of circadian rhythmicity in patients with moderate or severe AD^1^. Concerning sleep duration, both LS and SS have been previously shown to be linked with the risk of dementia^24^ ^74 75^. A 17-year longitudinal study investigating sleep characteristics in 11, 247 old-aged Swedish individuals (>65 years at baseline) observed an association of short (6 ≤ hours) and extended (> 9 hours) time in bed compared to the remaining individuals with a higher incidence of dementia (HR 1.40, 95% CI 1.06, 1.85; HR 1.11, 95% CI 1.00, 1.24)^24^. Our results are in agreement with the previously published study^24^. Indeed, we observed a strong causal role of SS in predisposition to AD (OR 1.26, 95% CI 1.08, 1.46). However, our results need to be treated with caution as the association was lost after excluding overlapping UKB samples from the AD dataset, as demonstrated previously^37^. However, it is also possible that the association was lost due to decreased sample size, necessitating replication in larger AD datasets in the future.

Sleep disturbances are also frequently observed in patients with ALS. Our MR analysis also suggested a possible causal role of INS on ALS (OR 1.55, 95% CI 1.12, 2.14). A previous observational study demonstrated decreased sleep efficiency and fragmented sleep architecture in 59 patients with ALS^76^. Another study reported the presence of sleep disturbances in more than 2/3 rd of 40 patients with ALS. The study further reported a diagnosis of INS in 65% of patients^77^. These results are in consent with a previous study reporting a significantly higher prevalence of INS in 90 patients with motor neuron disease compared to 96 healthy controls (48.9% vs 31.3%, p = 0.014)^78^. In summary, reports of sleep disturbance among patients with ALS in small sample-sized observational studies and suggestive causal role of INS on ALS in the present study necessitates a need for conducting large-scale epidemiological studies.

Despite the consistent findings of excessive daytime sleepiness or altered sleep timings in patients with PD, our MR findings demonstrate an absence of any causal role of sleep-related traits on predisposition to PD^79^ ^80^. One possible explanation could be that dopaminergic treatment might have influenced the sleeping behavior in patients with PD as excessive daytime sleepiness is known to be one of the common side effects of dopaminergic treatment^81^. In such a scenario, causal analysis using biological markers of circadian rhythms such as core body temperature, cortisol, and melatonin rhythms might potentially shed light on the true relationship between sleep-related traits and PD.

We also failed to observe any causal association of sleep-related traits with MS, although sleep disturbances are common symptoms in MS^82^. However, it is believed that sleep disorders observed in patients with MS could be secondary causes of fatigue in MS^83^, a symptom that affects 9 of the 10 patients with MS^84^.

Among all NDDs, a high prevalence of pain has been observed in AD and PD populations^85^ ^86^. Assessment of pain in such patients of often challenging due to associated cognitive and motor impairments^87^. Nevertheless, the use of genetic instruments of pain from a general population shows that MCP does not play any causal role in AD and PD. A recent cross-sectional study investigating pain in 100 PD patients further showed that pain is more prevalent in advanced-stage PD patients than early-stage PD patients, suggesting pain to be a consequence of the disease rather than a cause^85^. However, pain is a broad concept, and the inconsistencies in the measurement of a number of available pain behavior rating scales often limit their application in clinical settings.

### Strengths and limitations of this study

We adopted a highly comprehensive approach involving the exploration of several sleep-related traits and pain with commonly prevalent neurodegenerative disorders. We further employed multiple MR methods, heterogeneity, and sensitivity analyses approach, including causal check in the reverse direction to confirm the reliability of the observed associations. The MR approach relies on three main assumptions. Firstly, the genetic instrument must be strongly associated with the risk factor of interest. All our instruments had high F-statistics, which could be attributed to selecting the instruments through large-scale GWAS studies. Secondly, the genetic instruments must influence the disease of interest only through the risk factor. We did not find any direct influence of the SNPs included as a part of the genetic instruments with any NDD. Thirdly, none of the investigated SNPs should confound the relationship between the risk factor and disease of interest. In practice, it may not be entirely possible to test this assumption because of two main reasons: firstly, our lack of complete understanding of potential confounders of association of sleep and pain-related traits with NDDs and, secondly, the existence of a high proportion of pleiotropic variants in the human genome. Nevertheless, we adopted a highly conservative approach by excluding all pleiotropic variants to check the reliability of our associations. We also used several modern and robust MR methods, including WME and MBE, which are believed to provide more reliable estimates in the presence of invalid or weak instruments. Another potential source of bias could be overlapping samples between GWAS datasets used to investigate the association of genetic instruments with the risk factor and that used for association with the disease of interest. We observed that only AD and PD datasets had incorporated UKB samples which were also employed by GWAS on sleep-related traits and pain. Henceforth, we also used previously published AD and PD datasets that did not include UKB samples to rule out weak instrument bias. Such an approach also helped us to check the reliability of associations in the adoption of different diagnostic criteria for AD and PD by UKB and respective NDD consortia (The International Genomics of Alzheimer’s Project (IGAP) and International Parkinson Disease Genomics Consortium (IPDGC)). Another potential limitation of the present study is that we could not use 23andMe data for the computation of causal estimates due to the lack of open accessibility. The dataset may have shed more insights into the observed relationships by allowing us to not only replicate our findings but also provide pooled estimates. Previous observational studies have further shown that the impact of sleep and pain-related traits may be dependent on the stage of neurodegeneration or severity of the NDD^2^. However, we could not conduct such a stratified analysis due to the non-availability of an individual-level dataset for respective NDDs. And lastly, pain is a highly complex trait, and the lack of genetic instruments specific for neuropathic and nociceptive pain may undermine the findings of the present study. The possibility of nociceptive pain confounding the causal relationship between neuropathic pain and neurodegeneration cannot be ruled out.

### Conclusions and future research

Using genetic data, we provide strong evidence that being an MP is a causal risk factor for AMD. There is a necessity for the conduct of large-scale epidemiological cohort studies to confirm our findings. Additional research is also required to understand the biological pathways underlying these associations, including causal analysis with biochemical makers of sleep and correlated traits associated with sleep.

### What is already known on this topic

- Patients with NDD commonly report dysfunction in circadian rhythms.
- Numerous observational studies have shown associations between various sleep-related traits and NDD.
- As it is not possible to randomize sleep-related traits and longitudinal studies are difficult to conduct, specifically in elderly populations due to associated morbidity and mortality, results are highly conflicting.

### What this study adds

- Being an MP is a causal risk factor for AMD.
- The study necessitates further research on conducting stratified analysis based on the progression or severity of AMD.

## Author contributions

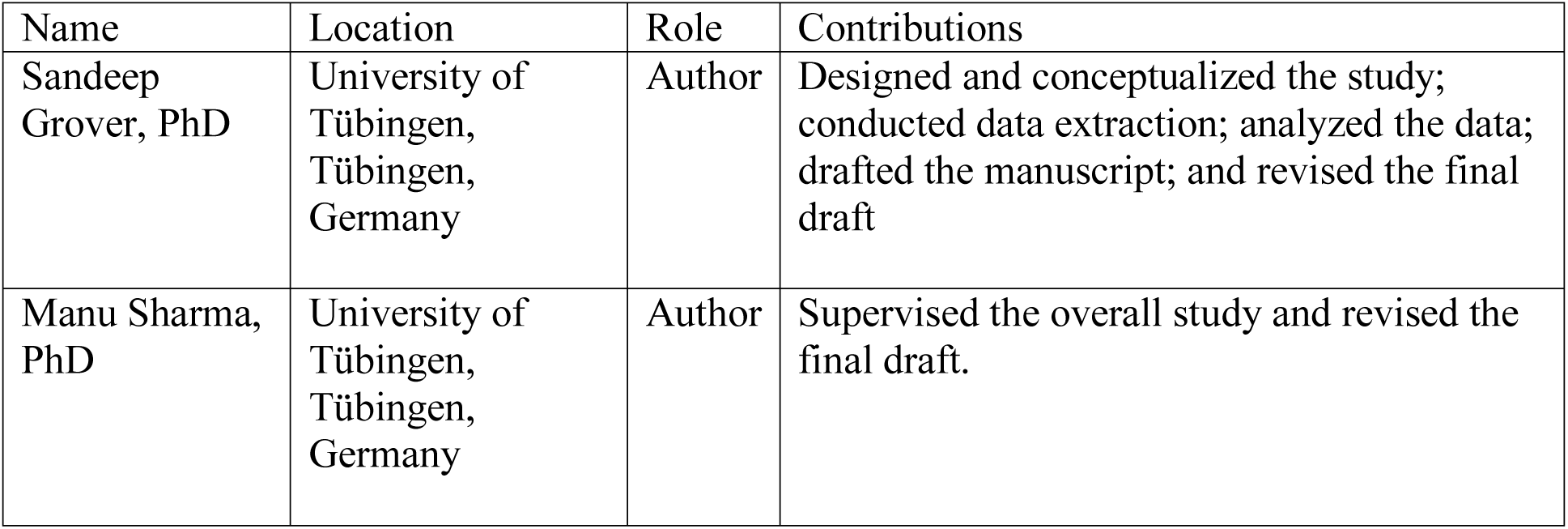

## Data Availability

All the data is publicly available.

## Acknowledgments

We acknowledge the study participants and investigators of International Genomics of Alzheimer’s patients (IGAP), International Parkinson’s Disease Genomics Consortium (IPDGC), project MinE, International Multiple Sclerosis Genetics Consortium (IMSGC), International Sleep Genetic Epidemiology Consortium (ISGEC), UK Biobank, UK Biobank sleep and chronotype research group for sharing the summary statistics on their GWAS datasets. We also acknowledge UK biobank resources under Application number 65949. This study is, in part, supported by the EU Joint Programme -Neurodegenerative Diseases Research (JPND) project under the aegis of JPND (www.jpnd.eu) through Germany, BMBF, funding code 01ED1406. M.S. is further funded by the Michael J Fox Foundation, USA Genetic Diversity in PD Program: GAP-India Grant ID: 17473. and also supported by the grants from the German Research Council (DFG/SH 599/6-1 to M.S.), MSA Coalition, and Michael J Fox Foundation.

---

**Supplementary Table 1.** Detectable effect size estimates for various neurodegenerative disorders (NDD) as an outcome.

**Supplementary Table 2.** Summary statistics used for the conduct of MR analysis in the present study.

**Supplementary Table 3.** Sensitivity analysis of causal effect estimated using leave-one-out SNP method

A. The causal effect of chronotype (CHR) on predisposition to age-related macular degeneration (AMD)

B. The causal effect of short sleep (SS) on predisposition to Alzheimer’s disease (AD)

C. The causal effect of insomnia (INS) on predisposition to amyotrophic lateral sclerosis (ALS)

